# Model-based Detection of Spatial Disease Boundaries Using Amortized Bayesian Inference

**DOI:** 10.64898/2026.06.21.26356187

**Authors:** Kyle Lin Wu, Sudipto Banerjee

## Abstract

Disease boundary analysis identifies abrupt changes in health outcomes across geographic boundaries, guiding targeted public health interventions and outbreak surveillance. Current implementations often adopt a Bayesian “wombling” approach and largely rely on Markov Chain Monte Carlo (MCMC) posterior sampling, presenting scalability issues for large-scale disease surveillance. We leverage amortized Bayesian inference (ABI) to accelerate the detection of spatial health disparities between neighboring US counties by embedding neural posterior estimation within a Bayesian areal wombling framework. Exploiting the computational efficiency of ABI, we further introduce the Residual Disparity Elimination Target, a metric for the required reduction in mortality or prevalence for a region to eliminate a significant disparity with its neighbor. We analyze tracheal, bronchus, and lung cancer mortality rates across mainland US counties and achieve results concordant with MCMC analysis while scaling areal wombling to hundreds of outcomes and translating disparity detection into interpretable policy objectives.

## 1. Introduction

Modern advances in computer science have fundamentally transformed spatial data analysis, giving rise to geospatial artificial intelligence (GeoAI), which integrates methods from machine learning and spatial science to analyze geographically referenced data. The applications of GeoAI span diverse geographical domains, including urban, transport, health, and environmental geography (Wang et al., 2024). In particular, deep learning techniques have enabled sophisticated analysis of environmental exposure (Iyer et al., 2025) alongside real-time disease surveillance and forecasting (Kamel Boulos et al., 2019). Despite the potential of deep learning to enhance data processing and predictive modeling in health disparity research (Abdel Magid et al., 2024), neural networks have seen limited application in spatial difference boundary analysis, the problem of detecting where a spatially varying outcome changes abruptly across geographic units or continuous space. Applications of boundary analysis include identifying local health inequities (Barboza-Salerno et al., 2025); studying social frontiers in human geography (Dean et al., 2019); defining ecological boundaries (Fitzpatrick et al., 2010); designing targeted public health interventions and sampling protocols across diverse health environments (Copeland, 2010; Jacquez, 2010); and identifying edge areas of disease or environmental exposure hot spots (Jacquez and Greiling, 2003a,b; Lee and Wen, 2023).

Established approaches to boundary analysis often adopt a statistical “wombling” framework, named for the pioneering work by Womble (1951). Recent probabilistic approaches for spatial data on continuous fields (e.g., temperature, pollution) include Banerjee and Gelfand (2006); Halder et al. (2024); Qu et al. (2021); and Quick et al. (2015). Approaches for areal models, which are our focus, include Aiello and Banerjee (2025); Corpas-Burgos and Martinez-Beneito (2020); Gao et al. (2023); Hanson et al. (2015); Lee and Mitchell (2012); Li et al. (2012, 2015); Lu et al. (2007); Ma et al. (2010); and Wu and Banerjee (2025). We focus particularly on areal models of geographically aggregated disease mortality or incidence rates. The aforementioned approaches adopt a Bayesian paradigm for inference, which quantifies uncertainty surrounding latent model parameters *θ* given the observed data *y* through the posterior probability distribution *p* (*θ* | *y*); such methods typically employ Markov Chain Monte Carlo (MCMC) methods (Brooks et al., 2011) to sample from an intractable posterior distribution. However, analyzing large collections of health outcomes via MCMC can become computationally expensive and difficult to scale, especially for non-parametric approaches, which limits their practicality for real-time surveillance applications.

To address these limitations, we draw on simulation-based inference (SBI; Cranmer et al., 2020), an alternative to MCMC that approximates Bayesian inference using simulated data from the joint model *p* (*y, θ*) without explicit evaluation of a likelihood function. Increasingly, SBI approaches employ neural networks to learn the relationship between observed data and latent parameters and enable amortized Bayesian inference (ABI), where the initial training period of the network accelerates subsequent inference to become orders of magnitude faster than gold-standard MCMC methods (Deistler et al., 2025; Kühmichel et al., 2026; Zammit-Mangion et al., 2025). The computational speedup of ABI most benefits repetitive inference tasks such as high-frequency disease surveillance across many health outcomes. We employ neural posterior estimation (NPE), a subfield of ABI (Zammit-Mangion et al., 2025, Section 3.2) that leverages generative models such as normalizing flows (Dinh et al., 2015; Kobyzev et al., 2021; Rezende and Mohamed, 2016; Tabak and Turner, 2013; Tabak and Vanden-Eijnden, 2010) for scalable, high-fidelity approximations to an intractable posterior *p* (*θ* | *y*) that retain full flexibility in posterior inference for boundary analysis. Alternative ABI approaches include neural Bayes estimation (Zammit-Mangion et al., 2025, Section 3.1), which trains neural networks to estimate posterior metrics (e.g., quantiles); neural likelihood estimation, which integrates a learned surrogate likelihood with standard inference methods such as MCMC (Lueckmann et al., 2019; Papamakarios et al., 2019); and neural posterior score estimation, which reverses a diffusion of the posterior distribution by learning the gradient of the log-density (Arruda et al., 2026a,b; Geffner et al., 2023; Sharrock et al., 2024).

Our work bridges GeoAI and boundary analysis by applying amortized Bayesian inference to detect spatial health disparities. We implement NPE to accelerate inference for any dataset of health outcomes over the US counties. In an analysis of tracheal, bronchus, and lung cancer mortality rates across the mainland US counties, we achieve estimation and boundary analysis results comparable to a standard MCMC approach under the areal wombling framework of Wu and Banerjee (2025). Furthermore, we utilize the computational advantages of NPE to derive data-driven health policy objectives from Bayesian areal wombling via a novel metric, the Residual Disparity Elimination Target (RDET), which represents the minimum reduction in disease morbidity or mortality that a higher residual risk county must achieve to eliminate the disparity with its neighbor after accounting for observed risk factors. By framing the inferential objective in terms of the health outcome, the RDET translates the results of Bayesian areal wombling into an actionable public health objective.

## 2. Results

We trained a neural posterior estimation network, hereby referred to as the NPE network, using the BayesFlow Python package (Kühmichel et al., 2026; Radev et al., 2022, 2023) to learn associations between any US county-level health outcome and several health risk factors under a Bayesian spatial regression model. Our model adjusted for county-level variations in smoking prevalence, physical inactivity, health insurance coverage, unemployment, as well as adult diabetes and obesity prevalence rates. We also incorporated the Social Vulnerability Index, a composite measure of socioeconomic status, household composition, demographics, housing, and transportation (Flanagan et al., 2011). By integrating this trained NPE network into a boundary analysis framework, we enabled rapid inference of county-level health disparities.

### 2.1. Tracheal, bronchus, and lung cancer mortality in the mainland US

We applied our analysis framework to investigate geographical disparities between neighboring US counties in age-standardized estimates of tracheal, bronchus, and lung cancer mortality rates in 2014 from the Institute of Health Metrics and Evaluation (Mokdad et al., 2017). Our analysis primarily considered spatial residual error representing disparities in mortality rates that are unexplained by the incorporated risk factors. Focusing on disparities between geographically adjacent counties, we subset our study region to 3,103 counties in the contiguous mainland United States with available risk factor data, containing 8,724 geographic boundaries between neighboring counties.

Statistically significant predictors of tracheal, bronchus, and lung cancer mortality, defined as variables with 95% credible intervals excluding zero, included smoking prevalence, social vulnerability, physical inactivity, and the uninsured rate (Table 1). Smoking prevalence had the largest standardized regression coefficient estimate, indicating the strongest association with mortality rates. Social vulnerability was moderately associated with higher mortality rates, while physical inactivity showed a weaker but significant positive association.

**Table 1.** Standardized regression coefficient estimates under a Bayesian spatial regression model predicting 2014 US county-level tracheal, bronchus, and lung cancer mortality rates. Estimates were obtained as the posterior mean of samples generated via a neural posterior estimation network, and 95% credible intervals were derived from the 2.5% and 97.5% posterior quantiles for regression coefficient.

| Variable | Posterior Mean | 95% Credible Interval |
| --- | --- | --- |
| Intercept | 0.010 | [-0.100, 0.122] |
| Smoking Prevalence | 0.424 | [0.389, 0.460] |
| Unemployment | 0.025 | [-0.015, 0.065] |
| Social Vulnerability Index | 0.104 | [0.055, 0.154] |
| Physical Inactivity | 0.060 | [0.029, 0.092] |
| Uninsured Rate | -0.166 | [-0.207, -0.125] |
| Diabetes Prevalence | 0.018 | [-0.011, 0.049] |
| Obesity Prevalence | 0.013 | [-0.016, 0.041] |

Having adjusted for the aforementioned risk factors, we identified 351 (4.02%) of the 8,724 total boundaries as significant residual disparities (Figure 1) using a Bayesian areal wombling approach to boundary detection (Wu and Banerjee, 2025). A spatial disparity between neighboring counties is identified by first computing the posterior probability of the standardized difference in residual mortality rates exceeding a specified difference threshold after adjusting for risk factors; we refer to this posterior metric as a “difference” probability. When the difference probability exceeds a defined significance threshold, the corresponding county-pair is reported as a spatial disparity. Both the difference and significance thresholds are automatically selected based on objective data criterion (Section 4.4).

**Figure 1.**
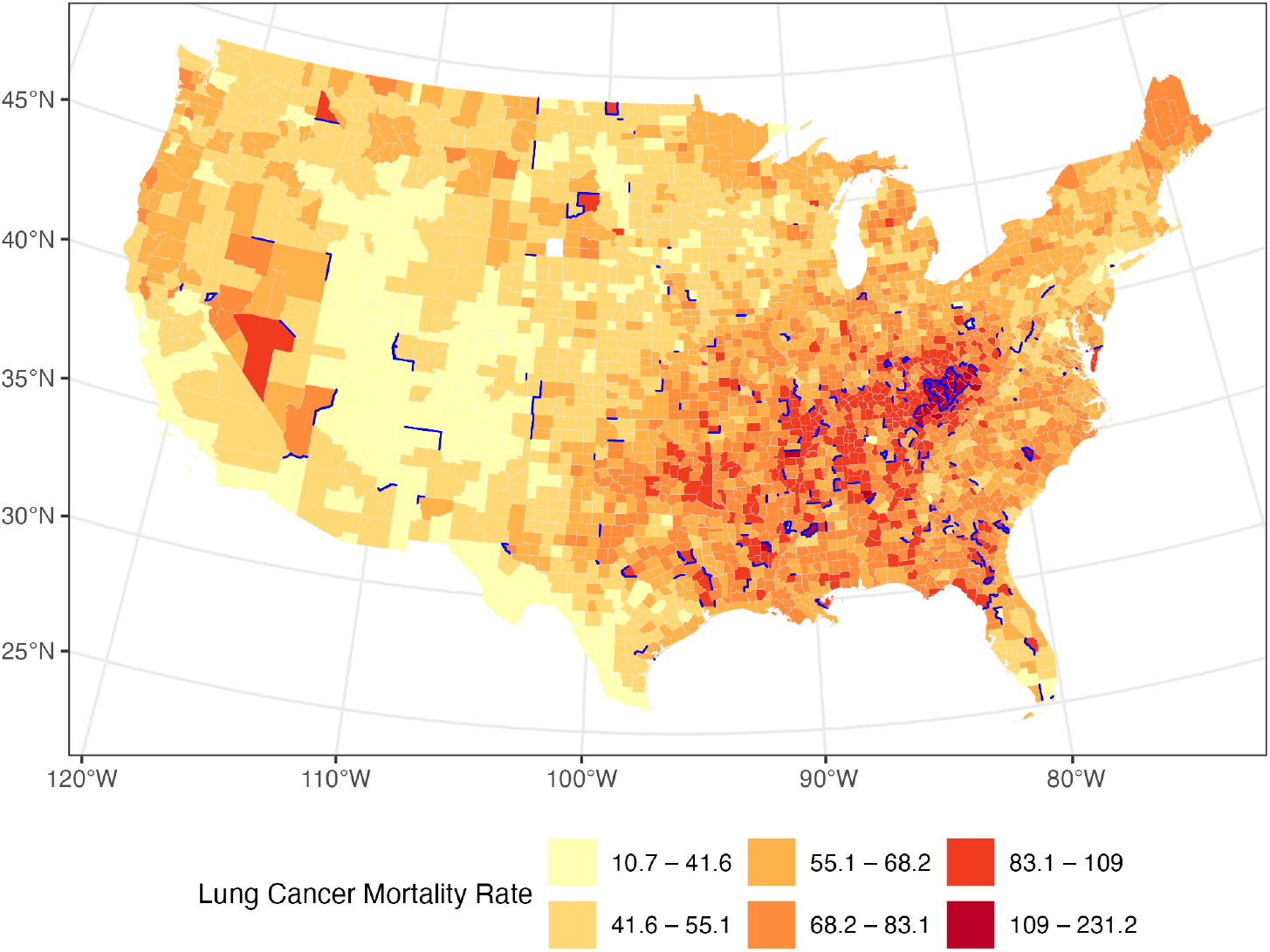
Mainland US county estimates of 2014 age-standardized tracheal, bronchus, and lung cancer mortality rates (counts per 100,000 population). Identified residual disparities are highlighted in blue. Disparity detection was performed using the Bayesian areal wombling framework proposed by Wu and Banerjee (2025) and accounted for county-level risk factors including smoking prevalence, physical inactivity, proportion of residents without health insurance, county social vulnerability index, adult diabetes prevalence, and adult obesity prevalence.

In Bayesian areal wombling, posterior difference probabilities provide uncertainty quantification for detecting spatial disparities. However, we instead sought an interpretable measure of disparity magnitude. To this end, we propose the Residual Disparity Elimination Target (RDET). The RDET for an identified disparity represents the minimum reduction in the mortality rate a county with a higher residual mortality rate must achieve to eliminate the significant health gap with its healthier neighbor, accounting for existing differences in risk factors. Within the 351 identified disparities between neighboring counties, we found 237 unique counties with higher residual mortality compared to neighboring counties, owing to the fact that multiple disparities may share the same county with a higher spatial residual risk. For each higher residual mortality county, we mapped the RDET in Figure 2; for counties identified as having a disparity with multiple neighboring counties, we mapped the minimum RDET across all reported disparities.

**Figure 2.**
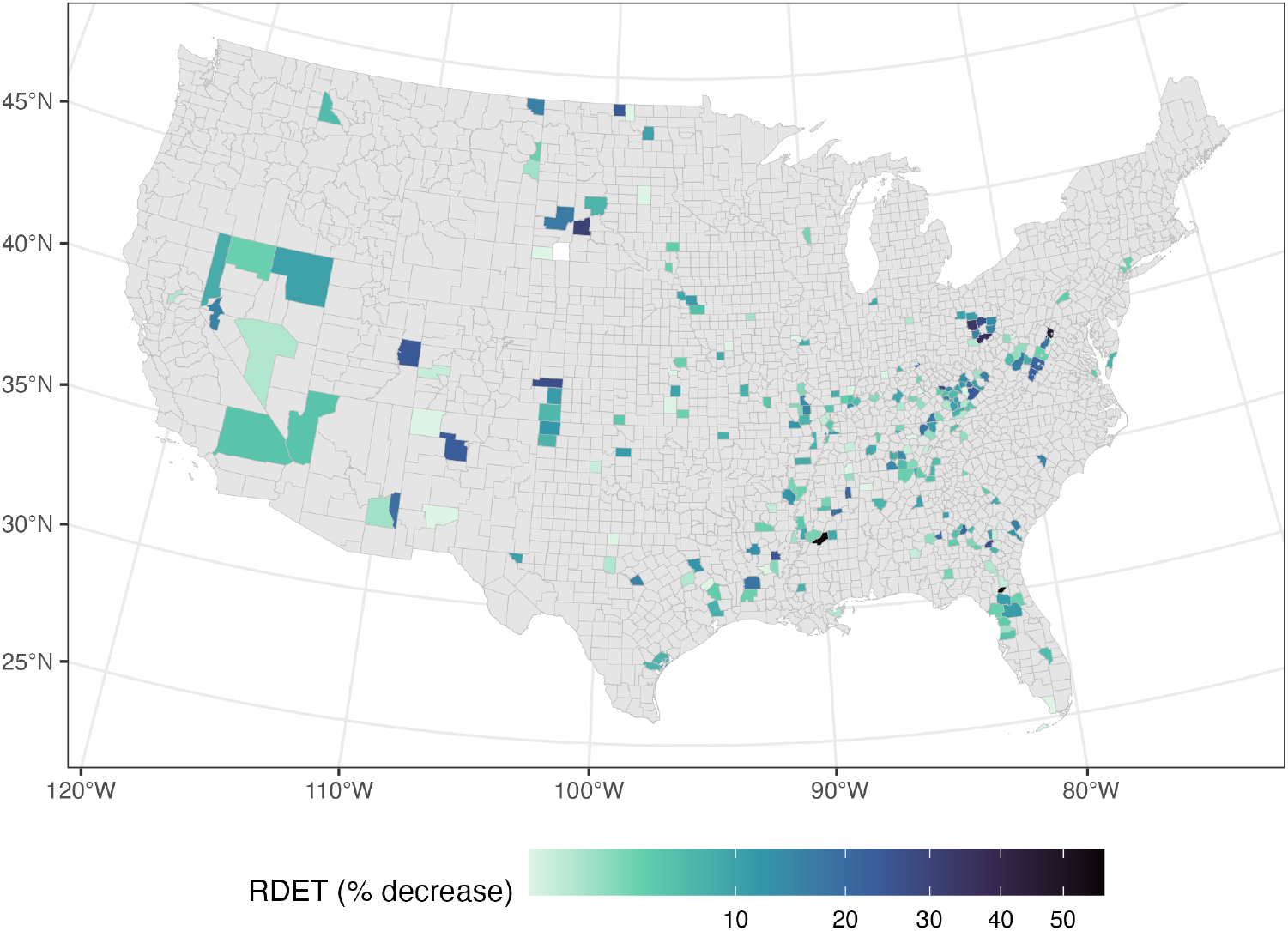
Residual disparity elimination targets (RDET) mapped as percent decrease of observed mortality rate for 237 US counties identified as higher residual mortality counties. The RDET for a detected disparity represents the minimum reduction in mortality rate at which a higher residual mortality county would not be classified as a disparity with a neighboring county, using the trained neural posterior estimator. Counties not identified as higher residual mortality counties are colored in gray.

Madison County, Mississippi, required the largest relative RDET reduction (57%, corresponding to a target age-standardized mortality rate of 55.6 per 100,000 population), representing the most substantial relative gap in estimated mortality compared with its neighboring counties after adjusting for risk factors (Table 2). In contrast, Union County, Florida, had a significantly higher RDET that represents a larger mortality rate but a smaller percentage decrease from the observed rate, resulting from differences in observed and latent risk factors in the surrounding regions. The full list of reported disparities and estimated RDETs is available in Table S1 in the supplementary material.

**Table 2.** Counties with ten largest Residual Disparity Elimination Target (RDET) percent reductions, estimated to the nearest half percent reduction. The annual mortality rate per 100,000 population corresponding to each RDET is listed and the percent decrease from the observed mortality is enclosed in parentheses. The reported RDET is the minimum value across all disparities for a higher residual mortality county.

| Higher Residual Mortality County | RDET (% decrease) |
| --- | --- |
| Madison County, Mississippi | 55.6 (57%) |
| Union County, Florida | 100.6 (56.5%) |
| Frederick County, Virginia | 36.7 (45.5%) |
| Washington County, Ohio | 47.7 (36.5%) |
| Muskingum County, Ohio | 56.8 (33.5%) |
| Haakon County, South Dakota | 39.4 (30.5%) |
| Powell County, Kentucky | 107.8 (28.5%) |
| Guernsey County, Ohio | 60.1 (27%) |
| Jeff Davis County, Georgia | 70.4 (26.5%) |
| Kiowa County, Colorado | 35.8 (26.5%) |

### 2.2. Comparison to Markov Chain Monte Carlo posterior sampling

We compared the speed of posterior sampling via the NPE network against Hamiltonian Monte Carlo (HMC) sampling, a specialized MCMC approach based on Hamiltonian mechanics, by implementing an equivalent analysis using the rstan R package (Stan Development Team, 2025). On a device equipped with an Apple M2 chip and 8 GB of RAM, the NPE network generated 8,000 posterior samples of regression coefficients and variance parameters for 200 simulated datasets in approximately 93 seconds. In contrast, using HMC on the same device to obtain 2, 000 × 4 chains = 8, 000 samples (after 1,000 burn-in samples per chain) for one dataset took ≈ 46 seconds. Scaling this baseline to 200 datasets without additional parallelization would require approximately 2.6 hours. Therefore, the NPE network offers a significant computational speedup by orders of magnitude, though this advantage comes at the cost of an initial training phase (Section 4.3). Therefore, amortized inference enables scalable inference for high-frequency disease surveillance applications or estimation of inferential quantities such as the RDET, which require Bayesian inference over many datasets.

We assessed the sensitivity of our results to the use of the neural posterior estimator and the choice of prior distribution by comparing the disparity detection results for the county-level tracheal, bronchus, and lung cancer mortality rates using the NPE network against two sets of analysis using HMC. We obtained two sets of posterior samples via HMC: one using a prior identical to the one used to train the NPE network, and one with a non-informative prior on the regression coefficients and variance parameters. Each set contained 2, 000 × 4 chains = 8, 000 samples after 1,000 burn-in samples per chain. We then repeated the boundary analysis with each set of HMC samples. The effective sample size was above 5,000, and the Gelman-Rubin statistic 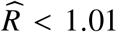 for all sampled parameters indicated reliable sampling from the true posteriors. The 95% credible intervals for the regression coefficients and variance parameters closely aligned for all three methods (Figure 3), indicating high quality posterior approximation and robustness with respect to the choice of prior distribution on the regression coefficients and variance parameters. Diabetes prevalence was marginally significant in the MCMC analyses but was insignificant when using the NPE network. The NPE network also slightly overestimated the total error variance (*σ*^2^) and the spatial proportion of variance (*ρ*) compared to the MCMC samples.

**Figure 3.**
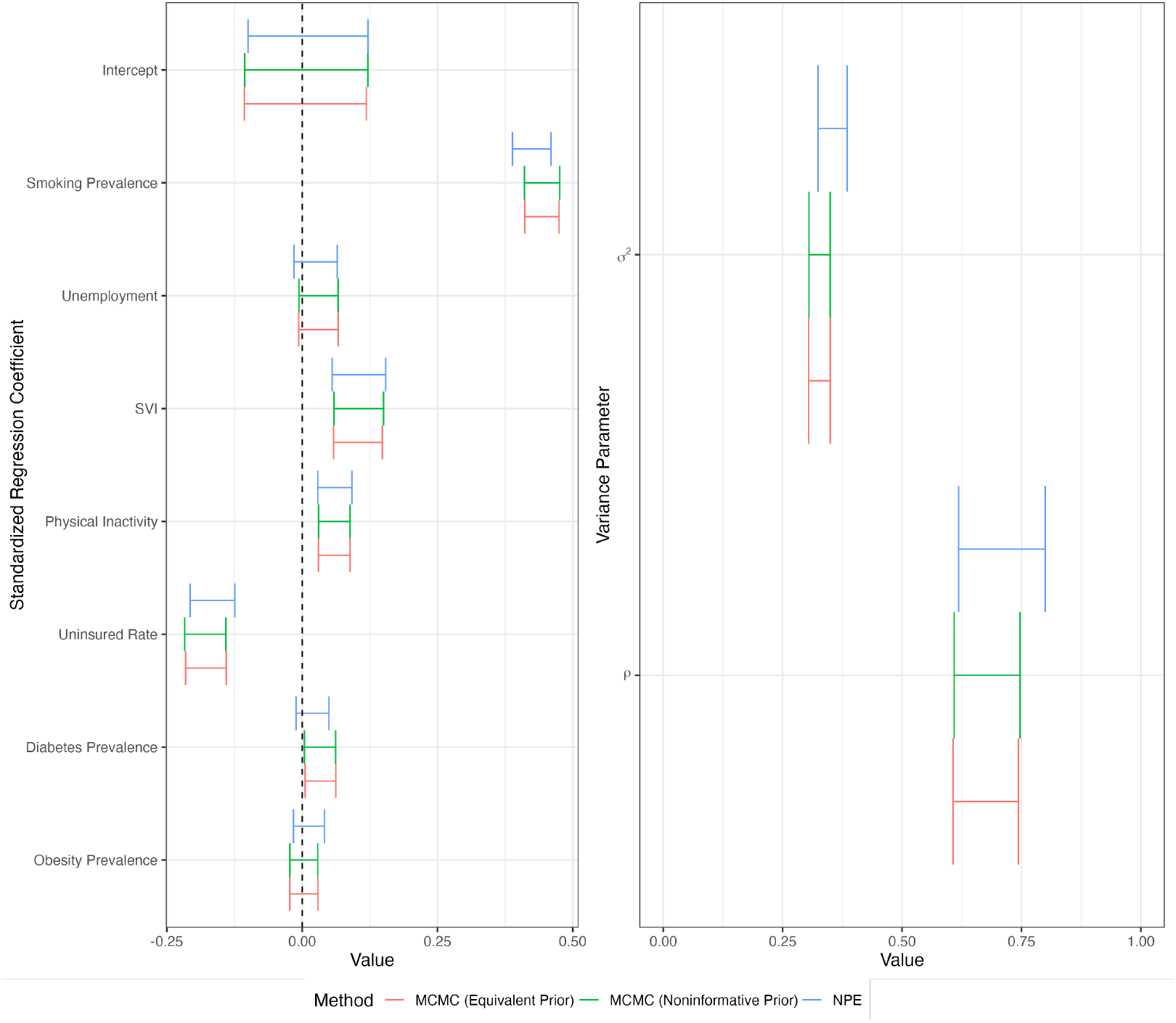
95% credible intervals of regression coefficients (left) and variance parameters (right) under a Bayesian spatial regression model of 2014 US county-level tracheal, bronchus, and lung cancer mortality rates, obtained from the neural posterior estimation (NPE) network, MCMC with an equivalent prior to the one used to train the NPE network, and MCMC with a non-informative prior on regression coefficients and variance parameters.

Turning to boundary detection, the posterior difference probabilities from the NPE network were highly correlated (*r* > 0.99) with those resulting from each MCMC approach analysis (Figure 4). Disparity classification was slightly more conservative when using MCMC with an equivalent prior, which resulted in reporting 328 (3.76%) out of 8,724 boundaries as disparities.

**Figure 4.**
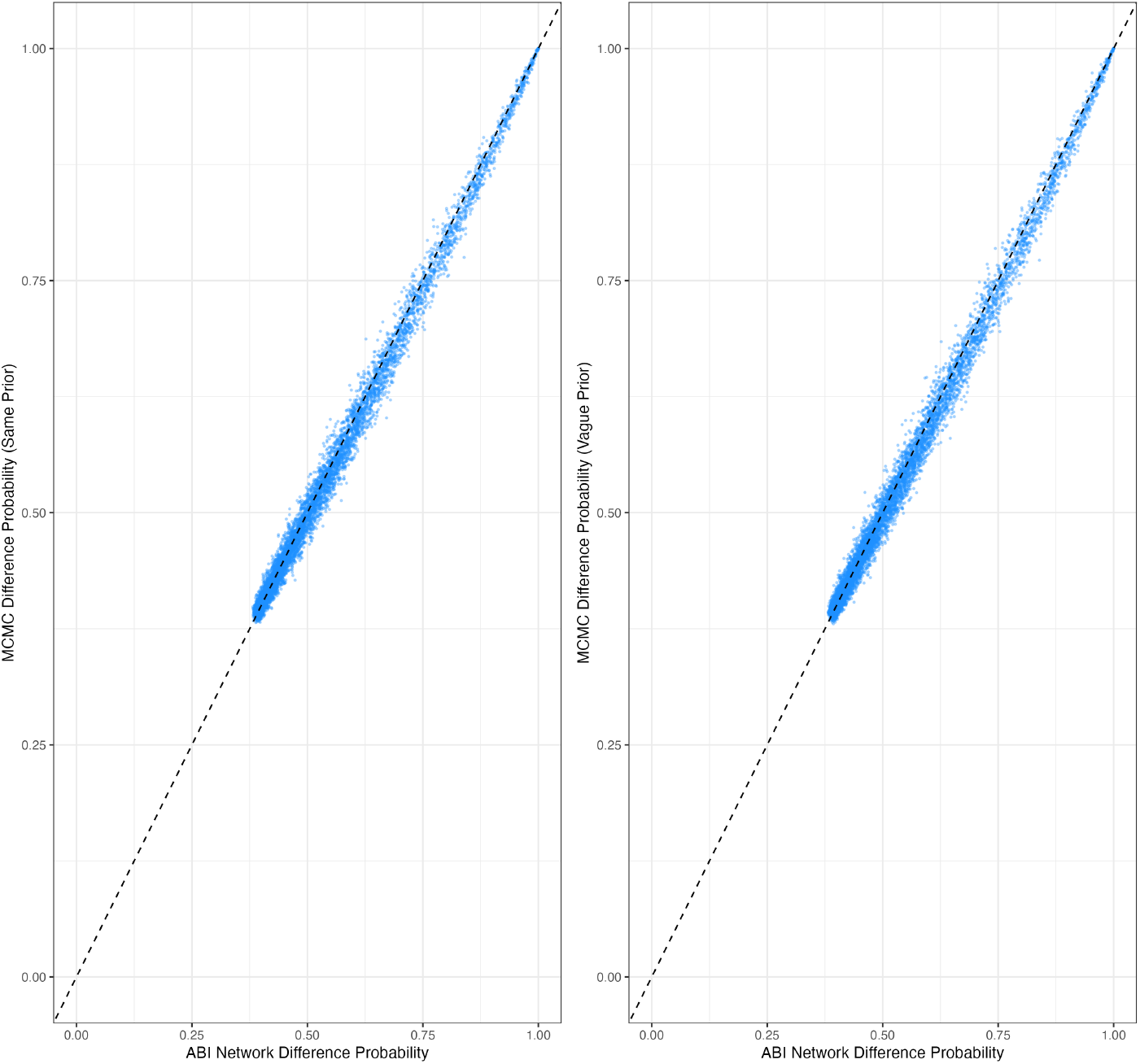
Neighboring county-pairs’ residual difference probabilities obtained using Markov Chain Monte Carlo (MCMC) with an equivalent prior (left) and non-informative prior (right) versus difference probabilities obtained using a neural posterior estimator. Higher difference probabilities indicate stronger evidence of a disparity between a pair of counties. The 45-degree reference line (dotted) denotes perfect equality.

## 3. Discussion

We introduce an AI-powered inference framework based on neural posterior estimation that is capable of rapid disparity detection, providing an objective assessment of local inequity in health outcomes. Our approach is well-suited for analyzing a large number of health outcomes or evaluating counterfactual scenarios that would be prohibitively computationally demanding using standard MCMC techniques. As a demonstration, we determined the reduction in specific counties’ mortality rates needed to eliminate statistical disparities with their neighboring counties. Thus, ABI allows for the efficient resolution of difficult inferential questions and offers a highly scalable tool for health inequity analysis, public health planning, and targeted interventions.

However, several limitations currently hinder the applicability of our approach for broad public health modeling or disease surveillance. First, our NPE network was trained with a fixed spatial adjacency structure and a set of risk factors. As a result, the network takes as input a complete dataset of health outcomes over the mainland US and would require retraining or data imputation if counties are missing data due to data suppression or reporting lag. Second, our NPE network is currently restricted to our specified spatial regression model, whose assumptions of normally-distributed outcomes and linear covariate effects may not be realistic. Third, approximating the full posterior of large-scale hierarchical models remains a challenge for ABI, which may compromise the accuracy of boundary detection in non-Gaussian models where the full conditional distribution of the random effects is not available.

Emerging methods such as compositional amortized inference with diffusion models (Arruda et al., 2026b) may provide improved performance in such settings. Finally, as with any analysis based on county-level data, our approach is subject to ecological fallacy and model misspecification effects, and detection biases may affect the validity of conclusions; the counterintuitive negative association between the uninsured rate and lung cancer mortality in our analysis may reflect surveillance bias or confounding by other variables such as rural-urban composition.

Potential future directions include expanding our NPE approach and boundary analysis framework to conduct inference for arbitrary maps and risk factor data, or employing more flexible simulators to account for richer dependencies across space, time, or multiple outcomes. We also foresee future explorations that adapt the supervised DeepRV framework of Navott et al. (2026) in probabilistic programming environments (Carpenter et al., 2017) to train our models. Alternatively, we can supervise any deep network using any method that delivers rapid posterior inference, as has been done by Presicce and Banerjee (2024) using Bayesian predictive stacking (Pan et al., 2025; Zhang et al., 2025), which avoids computationally expensive iterative algorithms such as MCMC.

## 4. Methods

### 4.1. Data sources and descriptions

Age-standardized tracheal, bronchus, and lung cancer mortality rate estimates in mainland US counties during 2014 were sourced from a publicly available dataset from the Institute of Health Metrics and Evaluation (Mokdad et al., 2017). US county-level estimates of 2014 county-level physical inactivity, adult diabetes prevalence, and adult obesity prevalence were extracted from the US Diabetes Surveillance System (Centers for Disease Control and Prevention). Additional covariates extracted from this source include 2014 unemployment and overall social vulnerability index percentiles (Flanagan et al., 2011), as well as the five-year average (2012-2016) of the proportion of uninsured residents aged 18 or older. Smoking prevalence estimates were sourced from publicly available IHME data, originally derived from the Behavioral Risk Factor Surveillance System (Dwyer-Lindgren et al., 2014).

We standardized the response and covariate data to have sample mean equal to zero and sample variance equal to one before statistical modeling. Since our analysis identifies spatial health disparities between neighboring counties, we restricted our study region to *n* = 3,103 mainland US counties with complete covariate and response data. Oglala Lakota County (formerly known as Shannon County), South Dakota, was excluded due to missing diabetes prevalence data. For subsequent spatial analysis, we determined adjacency relationships via rook contiguity in the 2014 cartographic boundary shapefile (U.S. Census Bureau, 2015).

### 4.2. Bayesian spatial regression model

We trained the NPE network on data simulated from a hierarchical spatial regression model. Following Wu and Banerjee (2025), we modeled associations between county-level health outcomes and risk factors using the “BYM2” model proposed by Riebler et al. (2016) as a reparameterization of the “BYM” model introduced by Besag et al. (1991). Let *y* denote an *n* × 1 vector *y* of health outcomes and *X* denote an *n* × (*p* + 1) design matrix containing an intercept column and *p* covariates. We modeled *y* using the BYM2 model,

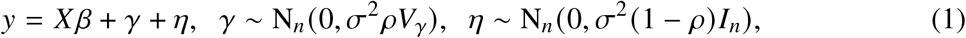

where *β* is a (*p* + 1) × 1 vector of intercept and regression coefficients, *γ* is an *n* × 1 vector of spatial effects, *η* is an *n* × 1 non-spatial error vector, *σ*^2^ > 0 represents the total error variance, *ρ* ∈ (0, 1) controls the proportion of spatial variance, and *V*_*γ*_ is a fixed positive definite *n* × *n* matrix encoding the spatial dependence structure. The spatial effects *γ* are of primary interest as representations of spatially autocorrelated latent factors driving local health inequity. We specified a conditionally autoregressive (CAR) prior for *γ* by defining the joint distribution of *γ* via conditional distributions of each spatial residual. We set *π*(*γ*_*i*_ | *σ*^2^, *ρ, γ*_−*i*_) = N(*α* ∑_*j, j*≠*i*_ *a*_*i j*_ *γ* _*j*_ /*a*_*i*+_, *cσ*^2^ *ρ*/*a*_*i*+_) for *i* = 1, …, *n* where *a*_*i j*_ is the binary adjacency indicator for counties *i* and *j, a*_*i*+_ = ∑_*j, j*≠*i*_ *a*_*i j*_ is the number of counties bordering county *i, α* is a spatial dependence parameter, and *c* is a fixed constant that scales the geometric mean of the prior marginal variances to one (Riebler et al., 2016). We fixed *α* = 0.99 to improve the identifiability of the spatial variance proportion parameter *ρ*. By Brook’s lemma (Chap. 4, Banerjee et al., 2015; Besag et al., 1991), the joint conditional prior is *π*(*γ* | *σ*^2^, *ρ*) = N_*n*_ (*γ* | 0, *σ*^2^*ρV*_*γ*_) as in (1), where 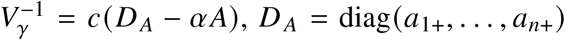, and *A* is the adjacency matrix.

We completed the model in (1) with a prior on *β, σ*^2^, and *ρ*. In ABI, the prior serves two purposes: encoding prior knowledge about the model parameters and determining the distribution of generated values for network training. Although non-informative priors are often deployed to minimize the prior’s influence on the posterior, overly diffuse priors may degrade posterior approximation quality due to simulated training datasets rarely resembling the observed data (Elsemüller et al., 2025; Frazier et al., 2024). In light of these considerations, we defined the complete joint prior as

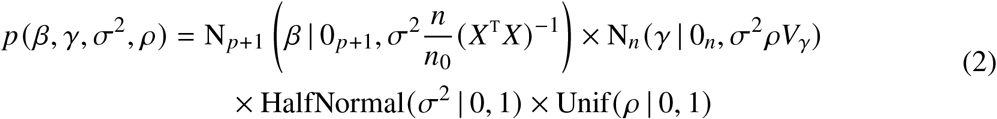

where the prior on *β* is a Zellner g-prior (Zellner, 1986) containing information equal to *n*_0_ = 5 independent observations. We assigned a semi-informative prior on *σ*^2^ equal to a standard Gaussian density truncated below zero since the residual error variance is unlikely to exceed 1.0 for standardized response and covariate data. In Section 2.2, we show that our results are not sensitive to this specification by performing an analysis using MCMC with a non-informative prior on the regression coefficients and total error variance.

To reduce the dimensionality of the target posterior, we focused on obtaining posterior samples of *β, σ*^2^ and *ρ*. To facilitate training, we reparameterized the three target parameters. First, we computed the QR factorization *X* = *QR*, where *Q* is an orthogonal matrix and *R* is upper-triangular, such that 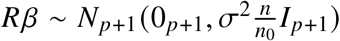. Second, to map each parameter space onto the unconstrained real line, we applied the log-transformation to *σ*^2^ and logistic transformation to *ρ* such that logit(*ρ*) = log (*ρ*/(1 − *ρ*)). Finally, we set the target parameters of the neural posterior estimator as *θ* = (*β*^T^ *R*^T^, log(*σ*^2^), logit(*ρ*))^T^. We employed the prior in (2) by fixing *X* using the risk factor data described in Section 4.1 to reduce training time and improve posterior approximation quality. To train the NPE network, we generated samples from *p* (*y, γ, θ*) by sampling (*β, γ, σ*^2^, *ρ*) from the prior in (2) and then drawing *y* ∼ *p* (*y* | *β, γ, σ*^2^, *ρ*) as in (1).

After the NPE network generated posterior samples of *θ*, posterior samples of the spatial effects *γ* (used for boundary detection in Section 4.4) were obtained via exact sampling from the full conditional,

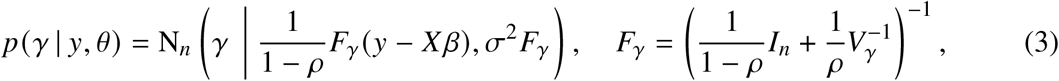

which can be done efficiently by pre-computing the eigen decomposition 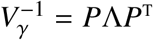, where *P* is an orthogonal *n*×*n* matrix and Λ is a diagonal matrix whose diagonal entries are the eigenvalues of 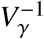. A sample of *γ* is obtained by drawing *z* ∼ N_*n*_ (0, *I*_*n*_) and taking *γ* = *S* (*S*^T^ (*y* − *X β*)/(1 − *ρ*) + *σz*), where *S* = *P* (1 − *ρ*)^−1^ *I*_*n*_ + *ρ*^−1^Λ ^−1/2^.

### 4.3. Amortized Bayesian inference

We detail our neural posterior estimation approach based on the framework by Radev et al. (2022) in Section 4.3.1 and describe our implementation in Section 4.3.2.

#### 4.3.1. Neural posterior estimation

Neural posterior estimation trains an “inference” network to sample the target posterior given arbitrary data, providing rapid approximate posterior inference. Here, the training data consists of simulated draws of the data and latent parameters from the models in (1) and (2). We focus on the implementation by Radev et al. (2022), where the inference network is a conditional invertible neural network (cINN) that learns a single-pass normalizing flow. Alternatively, recent developments in diffusion model approaches show potential for greater accuracy and training stability (Arruda et al., 2026a; Chen et al., 2025; Kühmichel et al., 2026).

In the framework proposed by Radev et al. (2022), an additional “summary” network preprocesses the observed data and outputs a learned summary vector, which the inference network then uses to define a mapping between a tractable base distribution and an intractable posterior. Although it is not strictly necessary for neural posterior estimation (see, for example, Zhou and Banerjee, 2026), the summary network can enable inference across datasets of varying size and perform feature extraction before feeding into the inference network. However, this may also result in information loss if the summary output is not sufficiently informative, particularly when the observations are non-exchangeable as in our spatial model. On the other hand, without a summary network, the inference network may struggle to extract sufficient information from the data and fail to recover the posterior distribution of the variance parameters.

Instead, we prevent information loss by passing both the response data *y* and a set of learned auxiliary features *h*_*ψ*_ (*y*) to the inference network, where *h*_*ψ*_ (·) is a customized auxiliary network with parameters *ψ* that extracts features using the adjacency structure of the map. Given the conditioning data, *y* and *h*_*ψ*_ (*y*), the feedforward process of the cINN defines a deterministic diffeomorphism from the parameter space to the latent base space. We denote this forward pass as the mapping *f*_*ϕ*_ (·; *y, h*_*ψ*_ (*y*)) : ℝ^*D*^ → ℝ^*D*^, where *ϕ* denotes the parameters of the inference network. Section S1 provides a detailed description of our auxiliary network.

In the training stage, the objective is network settings 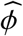 and 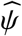 which lead to optimal approximation quality in the sense that they minimize the expected forward Kullback-Leibler divergence of the approximate posterior from the true posterior across all datasets *y*:

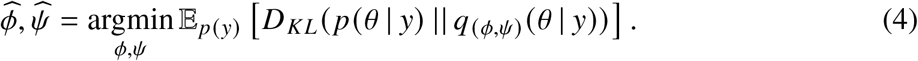

Here, *q*_(*ϕ,ψ*)_ (*θ* | *y*) is the probability distribution of a sample drawn via the NPE network from the approximate posterior of *θ* and the expectation is taken over the marginal density of the data, so minimizing (4) targets the approximation quality on average across all possible datasets. To accomplish the optimization in (4), stochastic optimization methods, such as Adam (Kingma and Ba, 2017), are employed to iteratively update network settings *ϕ* and *ψ* across generated batches of datasets. A batch of samples 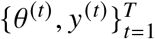 is generated by repeatedly sampling from the joint distribution *p* (*θ, y*) = *p* (*y* | *θ*) *p* (*θ*) specified by (1) and (2). Although we draw independent samples, active or sequential learning approaches may induce more efficient training (Durkan et al., 2018; Lueckmann et al., 2017; Papamakarios and Murray, 2018; Papamakarios et al., 2019).

The forward pass is necessary during training for tractability of the batch empirical loss given by

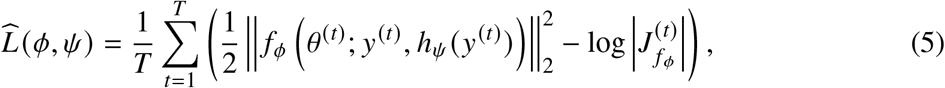

where 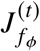 is the Jacobian matrix of *f*_*ϕ*_ (*θ*; *y*^(*t*)^, *h*_*ψ*_ (*y*^(*t*)^)) with respect to *θ*, evaluated at *θ* ^(*t*)^. Notably, this optimization in (4) and (5) can be executed without explicit evaluation of the likelihood or target posterior density and only requires sampling from *p* (*θ, y*). For the cINN architecture, the inverse function 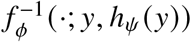 transforms samples from the base distribution, customarily a multivariate Gaussian, into samples from the target posterior. Thus, *q*_(*ϕ,ψ*)_ (*θ* | *y*) in (4) is the density of 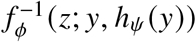, where *z* ∼ *N*_*D*_ (0, *I*_*D*_). Assuming perfect convergence (where the expected KL-divergence reaches 0), the distribution of generated samples is exactly the true posterior conditional on *y*. That is, 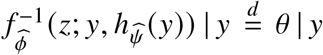 for all *y* in the support of *p* (*y*). Although perfect convergence is often not possible, high approximation quality can be achieved for a wide variety of datasets and validated using simulation-based calibration diagnostics (Modrák et al., 2025; Talts et al., 2018). The initial cost of training is amortized in subsequent inference sessions since sampling *z* and evaluation of 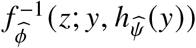 to obtain posterior samples of *θ* are both computationally cheap compared to MCMC approaches.

#### 4.3.2. Implementation for US Disease Maps

We constructed our inference network using the BayesFlow Python package with 8 sequential affine coupling layers (Dinh et al., 2017; Kingma and Dhariwal, 2018). For *i* = 1, …, 8, in layer *i*, the *D*-dimensional input vector *v*_*i*−1_ is passed through a permutation layer initialized once before training and fixed thereafter. The permuted vector *u*_*i*_ is then partitioned into *u*_*i*_ = (*u*_*i*1_, *u*_*i*2_), where *u*_*i*1_ contains the first ⌊*D*/2⌋ elements and *u*_*i*2_ contains the remaining ⌈*D*/2⌉ elements (⌊·⌋ and ⌈·⌉ denote floor and ceiling functions, respectively). The output *v*_*i*_ = (*v*_*i*1_, *v*_*i*2_) of layer *i* (and input for the subsequent layer) is

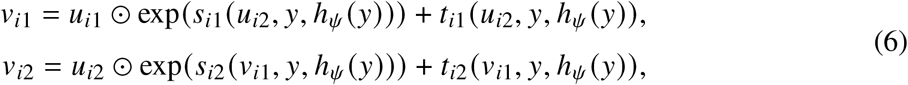

where ⊙ denotes element-wise multiplication, the exponential operator is applied element-wise, and *s*_*i*1_(·), *s*_*i*2_(·), *t*_*i*1_(·) and *t*_*i*2_(·) are each a multilayer perceptron comprising three hidden layers with 1,024 units and Swish activation functions (Ramachandran et al., 2017). For a training point (*θ* ^(*t*)^, *y*^(*t*)^), the first input vector is *v*_0_ = *θ* ^(*t*)^. The subnets *s*_*i*1_(·) and *t*_*i*1_(·) have ⌊*D*/2⌋-dimension output layers whereas *s*_*i*2_(·) and *t*_*i*2_(·) have ⌈*D*/2⌉-dimension output layers. The inference network parameters *ψ* is thus composed of the weights and biases of all subnets. After applying the forward pass in (6) sequentially, the final layer output is *v*_8_ = *f*_*ϕ*_ (*θ* ^(*t*)^; *y*^(*t*)^, *h*_*ψ*_ (*y*^(*t*)^)). The Jacobian of the affine coupling transformation is a triangular matrix, which allows cheap computation of the determinant term in the loss function of (5).

During the inference stage, samples of *z* are passed backward through the cINN. The inverse operation for layer *i* is given by

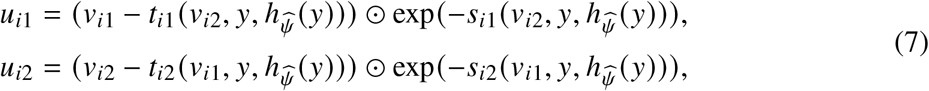

where the layer *i* permutation corresponding to *u*_*i*_ is inverted to obtain the input *v*_*i*−1_ for the next inverse layer. The final output 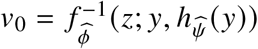 is a sample from 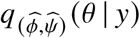. Similar to (6), the inverse pass in (7) also only requires evaluating the forward pass of each subnet, which enables computationally cheap sampling during inference. We obtained the network parameters 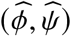 by training the inference and auxiliary networks with an Adam optimizer over 400 epochs; each epoch comprised 100 batches of 64 generated datasets. Training required 7.57 hours in Python v.3.11.13 using BayesFlow v.2.0.10 on a node of the Hoffman2 Shared Cluster at the University of California, Los Angeles (UCLA), utilizing 4 cores of an Intel Xeon Gold 6100 processor with 16 GB of total memory. Simulation-based calibration diagnostics in Section S2 of the supplementary material indicate excellent posterior calibration and recovery of the model parameters after training. Section S3 also shows that the inclusion of the auxiliary network improves performance compared to only passing the raw data *y* to the inference network.

To analyze the US county-level tracheal, bronchus, and lung cancer mortality rates, we generated 10,000 posterior samples of the global parameters *θ* = (*β*^T^ *R*^T^, log(*σ*^2^), logit(*ρ*))^T^ using the trained neural posterior estimator by drawing *z*^(*i*)^ ∼ *N*_*D*_ (0, *I*_*D*_) and setting 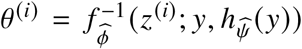 for *i* = 1, …, 10, 000. To obtain the corresponding posterior samples of *γ*, we drew one instance of *γ*^(*i*)^ ∼ *p* (*γ* | *y, θ* = *θ* ^(*i*)^) using (3) for *i* = 1, …, 10, 000. For each regression coefficient in *β*, the end points of the 95% credible intervals were estimated using the 2.5% and 97.5% posterior quantiles of the drawn samples. A county-level health risk factor was considered to be a significant predictor of tracheal, bronchus, and lung cancer mortality rate if the corresponding 95% credible interval excluded zero.

### 4.4. Boundary detection with Bayesian areal wombling

We adopted the Bayesian areal wombling approach to boundary detection proposed by Wu and Banerjee (2025) for assessing mortality rate disparities between all pairs of neighboring counties. We estimated difference probabilities of the form

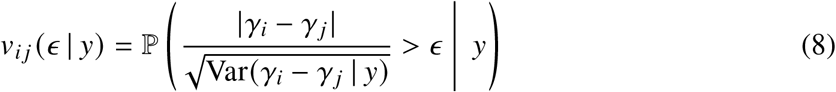

for all county pairs (*i, j*) such that *i* ∼ *j*, which denotes that county *i* borders county *j* and *i* < *j* for uniqueness. This difference probability represents the posterior evidence of a significant gap between the latent factors driving mortality rates between county *i* and *j* . The parameter *ϵ* > 0 is a difference threshold that is selected by minimizing the conditional entropy loss function,

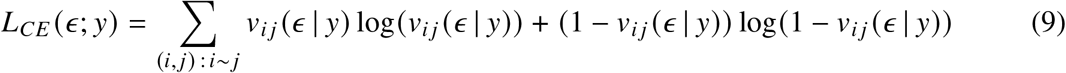

with respect to *ϵ* . To estimate the loss, we used posterior samples *γ*^(1)^, …, *γ*^(*S*)^ from the NPE network in Section 4.3 to substitute the Monte Carlo estimates 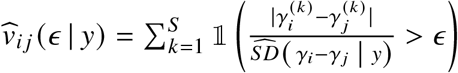 for *v*_*i j*_ in (9), where 1(·) denotes the indicator function and 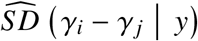 is the sample standard deviation of 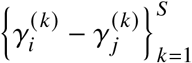. By minimizing (9), we obtained the optimal threshold as *ϵ*_★_ = 0.854 for the NPE analysis and *ϵ*_★_ = 0.849 for the baseline MCMC analysis described in Section 2.2. This process separates the most probable disparities from the remaining boundaries with ambiguous evidence and can be interpreted as regularizing against overconfidence in disparity classification. To estimate the conditional entropy loss, we substituted 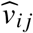 for *v*_*i j*_ in (9).

Next, we reported (*i, j*) as a disparity if 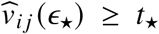 for a probability cutoff *t*_★_ ≥ 0 that we determined by controlling the Bayesian False Discovery Rate (BFDR) introduced in Muller et al. (2006). The BFDR quantifies the proportion of non-disparity boundaries expected to be misclassified as a disparity according to the posterior. Here, the BFDR for the chosen difference threshold *ϵ*_★_ is

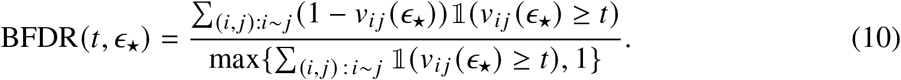

Given a pre-specified tolerance *δ* > 0, the corresponding difference probability cutoff is *t*_★_ = inf {*t* ∈ [0, 1] : BFDR(*t, ϵ*_★_) ≤ *δ*}, where the BFDR in (10) is estimated by substituting the posterior estimates 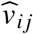 for *v*_*i j*_ . For the mortality rate analysis, we set *δ* = 0.05 and computed the cutoff as *t*_★_ = 0.895 for the NPE analysis and *t*_★_ = 0.897 for the baseline MCMC analysis in Section 2.2.

### 4.5. Residual Disparity Elimination Targets

We introduce the Residual Disparity Elimination Target (RDET) to provide a data-driven health policy objective that supplements disparity detection. The RDET can be adapted for any metric such as mortality, incidence, or prevalence; here, we use mortality for illustration. For a reported disparity (*i, j*), suppose that county *i* is the higher residual mortality county, i.e. E[*γ*_*i*_ − *γ* _*j*_ | *y*] > 0. Then, we define the RDET for disparity (*i, j*) as RDET(*i, j* ; *ϵ*_★_, *t*_★_) = sup{*y*_★_ ∈ ℝ : *v*_*i j*_ (*ϵ*_★_ | *y*_−*i*_, *y*_*i*_ = *y*_★_) < *t*_★_}, where *y*_−*i*_ denotes all observations except the *i*th region. Provided all risk factors and other county mortality rates remain constant, if county *i*’s mortality rate decreases to or below the RDET, the boundary detection method would no longer identify (*i, j*) as a significant disparity. Although an excessive decrease could invert the disparity classification such that county *i* becomes the low-residual risk, this scenario is not our primary focus.

We estimated RDET(*i, j* ; *ϵ*_★_, *t*_★_) for every disparity (*i, j*) using an iterative procedure. Without loss of generality, we assume county *i* is the higher residual mortality region. In each step *t* ≥ 0, we defined the reduction factor *c*_*t*_ = 1 − *ζ* (*t* + 1), where *ζ* is a pre-specified step size. In our implementation, we set *ζ* = 0.005 such that *c*_0_ = 0.995 in the first step. Let *x*_1_, …, *x*_*n*_ denote the unstandardized observed mortality rates, and recall that *y*_1_, …, *y*_*n*_ are the standardized mortality rates. At step *t*, we defined the hypothetical mortality rate 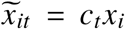. We then computed the standardized value 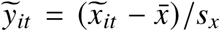 using the original mean 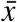 and standard deviation *s*_*x*_ of the observed county mortality rates. Next, we drew 3,000 samples of *θ* via the NPE network from 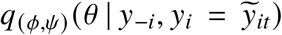 before drawing corresponding samples of *γ* from *p* (*γ* | *y, θ*). This step exploits the amortized network’s computational efficiency; unlike MCMC, which would require running separate chains for each hypothetical mortality rate, the network can draw samples for multiple time steps in seconds. We then implemented the boundary detection approach in Section 4.4 to estimate the counterfactual difference probability 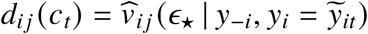. Finally, we set the estimate as 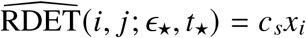, where *s* = min{*t* ∈ ℕ_0_ : *d*_*i j*_ (*c*_*t*_) < *t*_★_} is the final iteration corresponding to a (100 × (1 − *c*_*s*_))% decrease compared to the observed mortality *x*_*i*_.

## Data Availability

All data used in this paper are publicly available. Data on county-level estimates of physical inactivity, health insurance prevalence, unemployment, adult diabetes prevalence, adult obesity prevalence, and social vulnerability index analyzed during the current study are available from the US Diabetes Surveillance System at https://gis.cdc.gov/grasp/diabetes/diabetesatlas-analysis.html. The data on county-level tracheal, bronchus, and lung cancer mortality rates are available from the Institute for Health Metrics and Evaluation at https://ghdx.healthdata.org/record/ihme-data/united-states-cancer-mortality-rates-county-1980-2014. The data on county-level smoking prevalence estimates are available from the Institute for Health Metrics and Evaluation at https://ghdx.healthdata.org/record/ihme-data/united-states-smoking-prevalence-county-1996-2012.

## 6. Code Availability

All computer programs required to reproduce the data analysis in this manuscript are available at https://github.com/Ky-Wu/bayesflow_disparities.

## 7. Acknowledgments

This work used computational and storage services associated with the Hoffman2 Cluster which is operated by the UCLA Office of Advanced Research Computing’s Research Technology Group.

## 8. Author Contributions

KW and SB conceived the study. KW developed the code, carried out the data analysis, and wrote the initial draft of the manuscript. SB supervised the project and was a major contributor in editing the manuscript. All authors read and approved the final manuscript.

