## Supplementary Material for "Model-based Detection of Spatial Disease Boundaries Using Amortized Bayesian Inference"

KYLE LIN WU AND SUDIPTO BANERJEE

### S1. NPE Auxiliary Network Architecture

We detail the auxiliary network architecture used to generate additional conditioning statistics $h_\psi(y)$ for the NPE inference network in Section 4.3. Our approach computes learned auxiliary statistics from the ordinary least squares residuals and adjacency relationships. The learned auxiliary statistics $h_\psi(y)$ are concatenated with the original data $y$ and passed to the inference network. Although the auxiliary statistics do not increase the amount of theoretical information passed to the inference network, this feature extraction improves recovery of the variance parameters $\sigma^2$ and $\rho$ in the spatial regression model of (1), which we demonstrate in Section S3.

Let $y \in \mathbb{R}^n$ denote the observed responses, $X \in \mathbb{R}^{n\times(p+1)}$ denote the fixed design matrix with intercept and $p$ covariate vectors, and $A \in \mathbb{R}^{n\times n}$ denote the spatial adjacency matrix. During initialization, the network precomputes three matrices, (i) the projection matrix $P_{C(X)} = X(X^{\mathrm{T}}X)^{-1}X^{\mathrm{T}}$, (ii) the normalized adjacency matrix with self-loops $\widetilde{A}_{norm} = \widetilde{D}^{-1/2}\widetilde{A}\widetilde{D}^{-1/2}$ where $\widetilde{A} = A + I_n$ and $\widetilde{D}$ is a diagonal $n \times n$ matrix with diagonal elements $\widetilde{D}_{ii} = \sum_j \widetilde{A}_{ij}$, and (iii) the unnormalized graph Laplacian $L = D - A$, where $D$ is a diagonal $n \times n$ matrix with diagonal elements $D_{ii} = \sum_j A_{ij}$. During initialization, our auxiliary network also takes as arguments four constants, $d_1, d_2, d_3, d_4 \in \mathbb{N}$, that represent dimensions of hidden layer outputs detailed below.

First, the ordinary least squares residuals $r = (I_n - P_{C(X)})y$ are passed through a two-layer multi-layer perceptron (MLP) with Swish activation functions $\sigma_{Swish}(\cdot)$ to generate the feature representation $H = (h_1, \dots, h_n)^{\mathrm{T}} \in \mathbb{R}^{n\times d_1}$ with rows $h_i \in \mathbb{R}^{d_1}$ computed as

$$h_i = \sigma_{Swish}\left(W_2\sigma_{Swish}\left(w_1 r_i + b_1\right) + b_2\right)$$

for $i = 1, \dots, n$, where $w_1, b_1, b_2 \in \mathbb{R}^{d_1}$ and $W_2 \in \mathbb{R}^{d_1\times d_1}$. Next, the network captures spatial dependencies by computing three components based on the node representations and spatial adjacency: (i) a smooth component $S = \widetilde{A}_{norm}H$, (ii) a rough component $O = LH$, and (iii) a residual component $R = H - S$. These components are utilized to compute additional global spatial statistics. First, we compute the deviation statistics $\sigma_{nugget}, \tau_{nugget} \in \mathbb{R}^{d_1}$, which have elements given

*Date*: June 21, 2026.

by

$$\sigma_{nugget,j} = \sqrt{\frac{\sum_{i=1}^{n}(H_{ij} - \overline{H}_{\cdot j})^2}{n}}, \quad \tau_{nugget,j} = \sqrt{\frac{\sum_{i=1}^{n}(R_{ij} - \overline{R}_{\cdot j})^2}{n}},$$

for $j = 1, \ldots, d_1$, where $\overline{R}_{\cdot j}$ and $\overline{H}_{\cdot j}$ are the $j$th column means of $R$ and $H$, respectively. These statistics capture variation across regions in the learned features and residual components. We also compute the summary vector $Q \in \mathbb{R}^{d_1}$ where $Q_j$ is the Rayleigh quotient for the $j$th column in $H$,

$$Q_j = \frac{\sum_{i=1}^{n} H_{ij} O_{ij}}{\sum_{i=1}^{n} H_{ij}^2},$$

for $j = 1, \ldots, d_1$. Each statistic $Q_j$ measures the roughness or spatial variation of the $j$th latent feature. We then feed the extracted features and the smoothed component through another fully connected layer to obtain the compressed vector outputs

$$f = \sigma_{Swish}(W_c \text{vec}(H) + b_c), \quad f_s = \sigma_{Swish}(W_s \text{vec}(S) + b_s),$$

where vec$(\cdot)$ denotes the vectorization operator which vertically stacks the columns of a matrix, $W_c, W_s \in \mathbb{R}^{d_2 \times (nd_1)}$, and $b_c, b_s \in \mathbb{R}^{d_2}$.

Collecting the previous results, we concatenate the learned compressed features and global statistics into a $(3d_1 + 2d_2) \times 1$ summary vector $s = (f^{\mathrm{T}}, f_s^{\mathrm{T}}, Q^{\mathrm{T}}, \sigma_{nugget}^{\mathrm{T}}, \tau_{nugget}^{\mathrm{T}})^{\mathrm{T}}$. This summary vector is passed through another hidden layer and the final linear projection, yielding the learned auxiliary statistics

$$h_\psi(y) = W_{out}\sigma_{Swish}(W_h s + b_h) + b_{out},$$

where $W_{out} \in \mathbb{R}^{d_4 \times d_3}, W_h \in \mathbb{R}^{d_3 \times (3d_1 + 2d_2)}, b_h \in \mathbb{R}^{d_3}$, and $b_{out} \in \mathbb{R}^{d_4}$. During training, the learned parameters of the auxiliary network are $\psi = \{w_1, b_1, W_2, b_2, W_c, b_c, W_s, b_s, W_h, b_h, W_{out}, b_{out}\}$. For the trained auxiliary and inference networks used to generate the results in Section 2, we set $d_1 = 32, d_2 = 256, d_3 = 128$, and $d_4 = 64$.

### S2. NPE Simulation-based Calibration Diagnostics

We review simulation-based calibration diagnostics based on rank diagnostics for continuous variables (Modrák et al., 2025; Talts et al., 2018) and assess the trained NPE network presented in Section 2. Given a $D$-dimensional vector of continuous-valued parameters $\theta$, fix $j \in \{1, \ldots, D\}$. Let $F_{(\phi,\psi)}^{(j)}(\cdot \mid y)$ denote the cumulative density function (CDF) of the approximate posterior $q_{(\phi,\psi)}(\theta_j \mid y)$. If the posteriors match, i.e., $q_{(\phi,\psi)}(\theta_j \mid y) = p(\theta_j \mid y)$, and $x \sim p(\theta_j \mid y)$ is a draw from the true posterior, then $F_{(\phi,\psi)}^{(j)}(x) \sim \text{Unif}(0, 1)$. Given samples $\{y^{(t)}, \theta^{(t)}\}_{t=1}^{T}$ from the joint distribution $p(y, \theta)$, then the set of rank statistics $C_j = \left\{F_{(\phi,\psi)}^{(j)}\left(\theta_j^{(t)} \;\middle|\; y^{(t)}\right)\right\}_{i=1}^{T}$ is a set of i.i.d. draws from Unif$(0, 1)$ if the network is well-calibrated. For each draw, $F_{(\phi,\psi)}^{(j)}\left(\theta_j^{(t)} \;\middle|\; y^{(t)}\right)$ is estimated using posterior draws from the NPE network given data $y^{(t)}$. A significant difference between the empirical cumulative density function (ECDF) of $C_j$ and the CDF of Unif$(0, 1)$ indicates non-negligible approximation error for inference on $\theta_j$.

To assess the calibration of our trained NPE network, we simulated 500 datasets by sampling from the joint distribution $p(y, \beta, \sigma^2, \rho)$ specified by the likelihood and prior in Section 4.2 before using the trained NPE network to generate 2,000 posterior samples for each dataset. The resulting calibration diagnostics do not show significant departures from the theoretical cumulative density, suggesting a lack of significant evidence against convergence (Figure S1). The 95% credible intervals generated by the NPE also demonstrate excellent recovery of the global parameters generated from the prior (Figure S2).

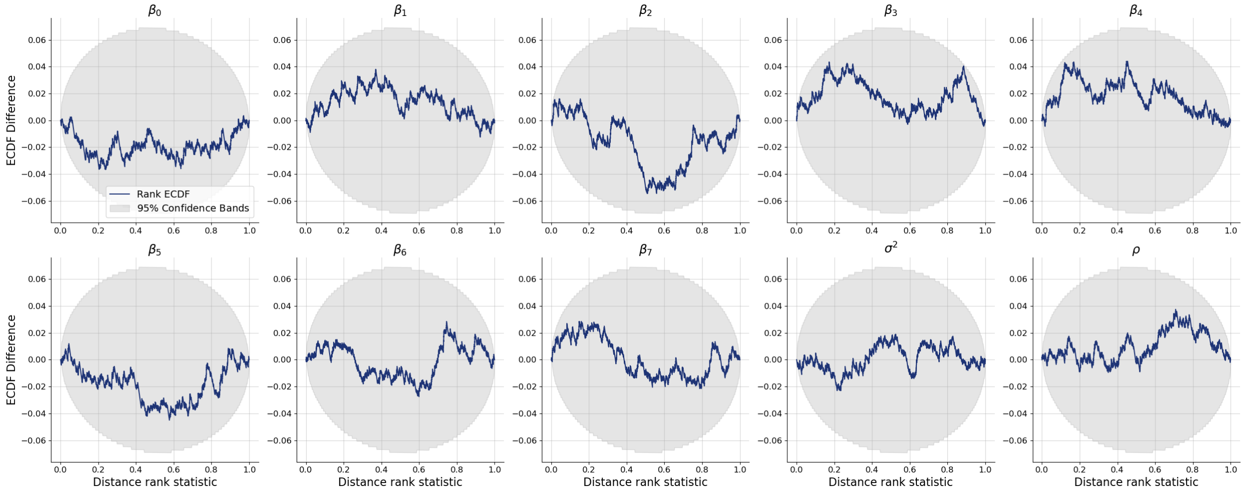


FIGURE S1. Simulation-based calibration plots of difference between empirical cumulative density function of simulated rank statistics and theoretical uniform distribution CDF. Grey bands indicate 95% confidence bands of the theoretical ECDF difference under convergence.

### S3. Impact of Auxiliary Network on Posterior Approximation Quality

As described in Section S1, we employ an auxiliary network to compute the learned statistics $h_\psi(y)$. We also pass the original response $y$ directly to the inference network, which departs from the dimension-reduction motivation for auxiliary networks proposed by Radev et al. (2022). Furthermore, since $h_\psi(y)$ is a function of $y$, concatenating them together theoretically does not increase the amount of available information. Nevertheless, the learned statistics $h_\psi(y)$ serve as extracted features that assist the inference network in learning about the variance parameters $\sigma^2$ and $\rho$. To evaluate the effectiveness of our customized auxiliary network, we fit a neural posterior estimator with the model and prior in Section 4.2 using the same training settings and inference network architecture as specified in Section 4.3 without the auxiliary network. Specifically, instead

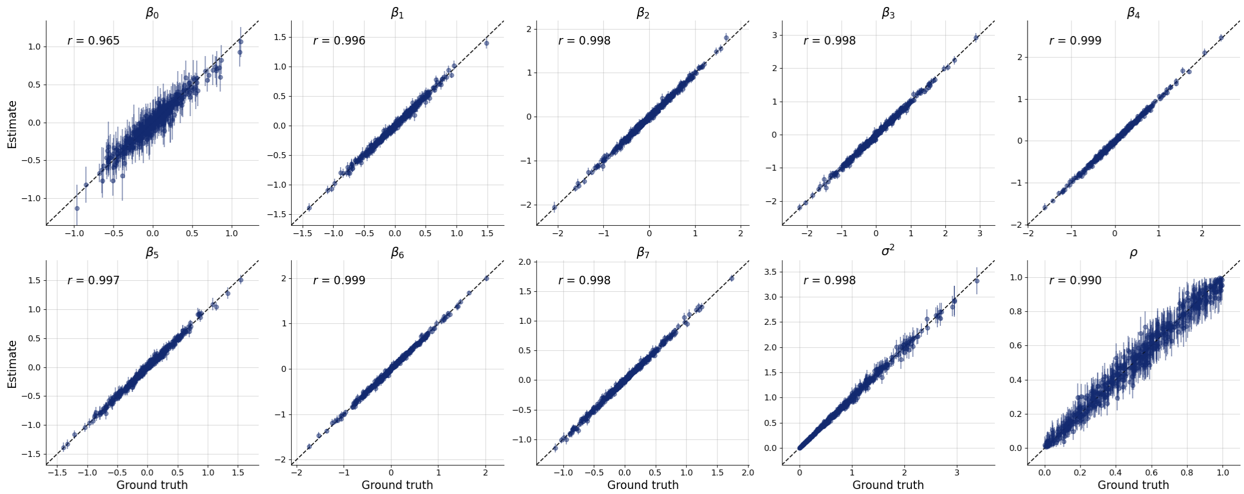


Figure S2. Recovery plot of credible intervals for regression coefficient and variance parameters generated by trained neural posterior estimator versus true values generated from the prior across 500 simulated datasets.

of passing $(y, h_\psi(y))$ as conditioning data to the inference network, we train an alternative network that only conditions on $y$.

After training, we simulated 500 datasets and used the alternative trained network to draw posterior samples for $\beta$, $\sigma^2$, and $\rho$, and re-conducted the comparison to a baseline MCMC approach as described in Section 2.2. Although the credible intervals show excellent recovery of the regression coefficients (similar to the results in Figure S2), the alternative network was significantly less precise in estimation of the variance parameters $\sigma^2$ and $\rho$ (Figure S3). This high inaccuracy also significantly impacts boundary detection, leading to large differences in the estimated difference probabilities compared to the baseline MCMC analysis (Figure S4).

### S4. Complete Set of Reported Disparities and RDETs

In Section 2.1, we detected significant differences in 2014 tracheal, bronchus, and lung cancer mortality rates between neighboring mainland US counties using Bayesian areal wombling and neural posterior estimation (Section 4). For each detected disparity, we computed the Residual Disparity Elimination Target (RDET), a metric quantifying the minimum reduction in the mortality rate of a county with higher residual mortality needed to eliminate its disparity with a neighboring county (Section 4.5). The complete list of detected disparities alongside their corresponding estimated difference probabilities and RDETs (computed as counts per 100,000 population and percent decrease from observed mortality rate) is listed in Table S1, ordered by descending RDET percent decreases.

| Higher Residual Mortality County | Lower Residual Mortality County | $\widehat{v}_{ij}(\epsilon_\star)$ | RDET (% decrease) |
|---|---|---|---|

| | | | |
|---|---|---|---|
| Madison County, Mississippi | Hinds County, Mississippi | 1.00 | 55.6 (57%) |
| Union County, Florida | Bradford County, Florida | 1.00 | 100.6 (56.5%) |
| Union County, Florida | Columbia County, Florida | 1.00 | 101.7 (56%) |
| Union County, Florida | Baker County, Florida | 1.00 | 102.9 (55.5%) |
| Union County, Florida | Alachua County, Florida | 1.00 | 106.4 (54%) |
| Madison County, Mississippi | Rankin County, Mississippi | 1.00 | 60.2 (53.5%) |
| Frederick County, Virginia | Hardy County, West Virginia | 1.00 | 36.7 (45.5%) |
| Madison County, Mississippi | Attala County, Mississippi | 1.00 | 72.5 (44%) |
| Madison County, Mississippi | Yazoo County, Mississippi | 1.00 | 73.1 (43.5%) |
| Washington County, Ohio | Noble County, Ohio | 1.00 | 47.7 (36.5%) |
| Madison County, Mississippi | Scott County, Mississippi | 1.00 | 82.8 (36%) |
| Muskingum County, Ohio | Noble County, Ohio | 1.00 | 56.8 (33.5%) |
| Haakon County, South Dakota | Ziebach County, South Dakota | 1.00 | 39.4 (30.5%) |
| Powell County, Kentucky | Montgomery County, Kentucky | 1.00 | 107.8 (28.5%) |
| Powell County, Kentucky | Clark County, Kentucky | 1.00 | 110.1 (27%) |
| Guernsey County, Ohio | Noble County, Ohio | 1.00 | 60.1 (27%) |
| Jeff Davis County, Georgia | Wheeler County, Georgia | 1.00 | 70.4 (26.5%) |
| Kiowa County, Colorado | Crowley County, Colorado | 0.98 | 35.8 (26.5%) |
| Caldwell Parish, Louisiana | Franklin Parish, Louisiana | 1.00 | 84 (25.5%) |
| Madison County, Mississippi | Leake County, Mississippi | 1.00 | 97 (25%) |
| Rolette County, North Dakota | Pierce County, North Dakota | 1.00 | 80.6 (24.5%) |
| Grand County, Utah | San Juan County, Utah | 0.99 | 41.1 (24%) |
| Pike County, Kentucky | Buchanan County, Virginia | 1.00 | 90.7 (23.5%) |
| Sandoval County, New Mexico | McKinley County, New Mexico | 0.97 | 26.3 (23.5%) |
| Rockbridge County, Virginia | Buena Vista City, Virginia | 0.97 | 42.6 (22%) |
| Augusta County, Virginia | Pendleton County, West Virginia | 0.98 | 47.2 (21%) |
| Houston County, Georgia | Dooly County, Georgia | 0.98 | 50.2 (20.5%) |
| Chattahoochee County, Georgia | Marion County, Georgia | 1.00 | 66.4 (20.5%) |
| Gallatin County, Kentucky | Boone County, Kentucky | 1.00 | 107.4 (20.5%) |
| Lee County, Mississippi | Union County, Mississippi | 1.00 | 84.8 (20.5%) |
| Greenlee County, Arizona | Apache County, Arizona | 0.97 | 38.2 (19%) |
| Webster County, Mississippi | Choctaw County, Mississippi | 0.99 | 74.1 (18.5%) |
| Powell County, Kentucky | Menifee County, Kentucky | 1.00 | 123.6 (18%) |
| Vernon Parish, Louisiana | Allen Parish, Louisiana | 0.99 | 71.5 (18%) |
| Fentress County, Tennessee | Cumberland County, Tennessee | 1.00 | 87.6 (18%) |
| Martin County, Kentucky | Wayne County, West Virginia | 1.00 | 102.7 (18%) |
| McCreary County, Kentucky | Wayne County, Kentucky | 1.00 | 118.4 (17.5%) |
| Shenandoah County, Virginia | Hardy County, West Virginia | 0.99 | 52.6 (17.5%) |
| Meade County, South Dakota | Ziebach County, South Dakota | 0.98 | 47.5 (17%) |
| Rolette County, North Dakota | Bottineau County, North Dakota | 1.00 | 89.2 (16.5%) |
| Pocahontas County, West Virginia | Pendleton County, West Virginia | 0.98 | 59 (16.5%) |

| | | | |
|---|---|---|---|
| Gallatin County, Kentucky | Carroll County, Kentucky | 1.00 | 113.5 (16%) |
| Sheridan County, Montana | Divide County, North Dakota | 0.98 | 55.8 (16%) |
| Morgan County, Ohio | Noble County, Ohio | 0.98 | 56.2 (16%) |
| Effingham County, Georgia | Jasper County, South Carolina | 0.99 | 65.4 (16%) |
| Gallatin County, Kentucky | Owen County, Kentucky | 1.00 | 114.2 (15.5%) |
| Lyon County, Nevada | Storey County, Nevada | 0.98 | 61.1 (15.5%) |
| Lampasas County, Texas | San Saba County, Texas | 0.99 | 72.1 (15.5%) |
| Boone County, West Virginia | Raleigh County, West Virginia | 1.00 | 100.7 (15.5%) |
| Gallatin County, Kentucky | Switzerland County, Indiana | 1.00 | 114.9 (15%) |
| Marlboro County, South Carolina | Robeson County, North Carolina | 0.99 | 104 (15%) |
| Monroe County, Ohio | Noble County, Ohio | 0.98 | 52.8 (15%) |
| Owsley County, Kentucky | Jackson County, Kentucky | 1.00 | 119.8 (14.5%) |
| Madison County, Missouri | Perry County, Missouri | 0.99 | 86 (14.5%) |
| Lincoln County, Tennessee | Moore County, Tennessee | 0.98 | 67.4 (14.5%) |
| Caldwell Parish, Louisiana | Ouachita Parish, Louisiana | 0.99 | 96.9 (14%) |
| Lee County, Mississippi | Pontotoc County, Mississippi | 0.99 | 91.8 (14%) |
| Saint Francois County, Missouri | Sainte Genevieve County, Missouri | 0.99 | 82.2 (14%) |
| Rolette County, North Dakota | Towner County, North Dakota | 1.00 | 91.8 (14%) |
| Belmont County, Ohio | Noble County, Ohio | 0.97 | 56.5 (14%) |
| Marlboro County, South Carolina | Florence County, South Carolina | 0.99 | 105.2 (14%) |
| Coffee County, Tennessee | Moore County, Tennessee | 0.98 | 68.4 (14%) |
| Jeff Davis County, Georgia | Telfair County, Georgia | 0.98 | 82.8 (13.5%) |
| Lawrence County, Kentucky | Boyd County, Kentucky | 0.99 | 105.9 (13.5%) |
| Bienville Parish, Louisiana | Red River Parish, Louisiana | 0.98 | 77.8 (13%) |
| Chattahoochee County, Georgia | Stewart County, Georgia | 0.98 | 73.1 (12.5%) |
| Hardin County, Illinois | Pope County, Illinois | 0.99 | 92.7 (12.5%) |
| Gallatin County, Kentucky | Grant County, Kentucky | 0.99 | 118.2 (12.5%) |
| McCreary County, Kentucky | Laurel County, Kentucky | 0.99 | 125.6 (12.5%) |
| Wolfe County, Kentucky | Morgan County, Kentucky | 0.99 | 103.5 (12.5%) |
| Arkansas County, Arkansas | Lincoln County, Arkansas | 0.98 | 72.3 (12.5%) |
| Frederick County, Virginia | Hampshire County, West Virginia | 0.97 | 58.9 (12.5%) |
| Bryan County, Georgia | Chatham County, Georgia | 0.98 | 74.4 (12%) |
| Carter County, Missouri | Oregon County, Missouri | 0.99 | 87.9 (12%) |
| Cimarron County, Oklahoma | Union County, New Mexico | 0.97 | 51.3 (12%) |
| Anderson County, Texas | Cherokee County, Texas | 0.99 | 92.7 (12%) |
| Fulton County, Kentucky | Hickman County, Kentucky | 0.99 | 93.1 (11.5%) |
| Johnson County, Kentucky | Morgan County, Kentucky | 0.99 | 111.6 (11.5%) |
| Marlboro County, South Carolina | Richmond County, North Carolina | 0.99 | 108.2 (11.5%) |
| Alachua County, Florida | Marion County, Florida | 0.95 | 51.8 (11%) |
| Marion County, Florida | Sumter County, Florida | 0.96 | 59.6 (11%) |
| Pike County, Georgia | Meriwether County, Georgia | 0.98 | 75.6 (11%) |

| | | | |
|---|---|---|---|
| Harrison County, Iowa | Crawford County, Iowa | 0.97 | 59.8 (11%) |
| Scott County, Tennessee | Wayne County, Kentucky | 0.99 | 111.6 (11%) |
| Dewey County, Oklahoma | Major County, Oklahoma | 0.97 | 66.1 (11%) |
| Marlboro County, South Carolina | Chesterfield County, South Carolina | 0.99 | 108.9 (11%) |
| Anderson County, Texas | Houston County, Texas | 0.99 | 93.8 (11%) |
| Prowers County, Colorado | Bent County, Colorado | 0.96 | 54.2 (11%) |
| Leslie County, Kentucky | Bell County, Kentucky | 0.99 | 126 (10.5%) |
| Perry County, Kentucky | Clay County, Kentucky | 0.99 | 133.3 (10.5%) |
| Coshocton County, Ohio | Holmes County, Ohio | 0.96 | 63.4 (10.5%) |
| Scott County, Tennessee | Pickett County, Tennessee | 0.98 | 112.3 (10.5%) |
| Loving County, Texas | Eddy County, New Mexico | 0.97 | 70.7 (10.5%) |
| Mingo County, West Virginia | Buchanan County, Virginia | 0.98 | 104.4 (10.5%) |
| Cleburne County, Alabama | Randolph County, Alabama | 0.97 | 81 (10%) |
| Montgomery County, Georgia | Wheeler County, Georgia | 0.97 | 71.3 (10%) |
| Chase County, Kansas | Marion County, Kansas | 0.97 | 67 (10%) |
| McCreary County, Kentucky | Pulaski County, Kentucky | 0.99 | 129.2 (10%) |
| Magoffin County, Kentucky | Morgan County, Kentucky | 0.99 | 103.2 (10%) |
| Lee County, Mississippi | Prentiss County, Mississippi | 0.98 | 96 (10%) |
| Nelson County, North Dakota | Benson County, North Dakota | 0.96 | 54.8 (10%) |
| Knox County, Ohio | Holmes County, Ohio | 0.95 | 52.4 (10%) |
| Scott County, Tennessee | Anderson County, Tennessee | 0.99 | 112.9 (10%) |
| Elko County, Nevada | Box Elder County, Utah | 0.94 | 53.8 (10%) |
| Hardin County, Kentucky | Bullitt County, Kentucky | 0.96 | 79.6 (9.5%) |
| Sharkey County, Mississippi | Issaquena County, Mississippi | 0.97 | 68.5 (9.5%) |
| Lee County, Mississippi | Monroe County, Mississippi | 0.97 | 96.6 (9.5%) |
| Burt County, Nebraska | Cuming County, Nebraska | 0.96 | 59 (9.5%) |
| Starke County, Indiana | Marshall County, Indiana | 0.98 | 93.3 (9%) |
| Leake County, Mississippi | Neshoba County, Mississippi | 0.97 | 82.3 (9%) |
| Clinton County, Missouri | DeKalb County, Missouri | 0.96 | 69.4 (9%) |
| Dade County, Georgia | Hamilton County, Tennessee | 0.98 | 82.9 (9%) |
| Hickman County, Tennessee | Williamson County, Tennessee | 0.98 | 96.4 (9%) |
| Aransas County, Texas | Nueces County, Texas | 0.96 | 64.7 (9%) |
| Richmond County, Georgia | Edgefield County, South Carolina | 0.95 | 68.6 (8.5%) |
| Cass County, Illinois | Brown County, Illinois | 0.96 | 70.2 (8.5%) |
| Franklin County, Illinois | Perry County, Illinois | 0.97 | 85.5 (8.5%) |
| Harrison County, Iowa | Shelby County, Iowa | 0.95 | 61.5 (8.5%) |
| Owsley County, Kentucky | Clay County, Kentucky | 0.99 | 128.3 (8.5%) |
| Carter County, Missouri | Reynolds County, Missouri | 0.97 | 91.4 (8.5%) |
| Washoe County, Nevada | Storey County, Nevada | 0.93 | 46.8 (8.5%) |
| Trousdale County, Tennessee | Sumner County, Tennessee | 0.98 | 93.7 (8.5%) |
| Lampasas County, Texas | Burnet County, Texas | 0.97 | 78.1 (8.5%) |

| | | | |
|---|---|---|---|
| Lampasas County, Texas | Mills County, Texas | 0.96 | 78.1 (8.5%) |
| Poinsett County, Arkansas | Craighead County, Arkansas | 0.98 | 102.6 (8.5%) |
| Highland County, Virginia | Pendleton County, West Virginia | 0.95 | 45.2 (8.5%) |
| Starke County, Indiana | Saint Joseph County, Indiana | 0.98 | 94.3 (8%) |
| Harlan County, Kentucky | Wise County, Virginia | 0.98 | 123.2 (8%) |
| Perry County, Kentucky | Knott County, Kentucky | 0.98 | 137.1 (8%) |
| East Feliciana Parish, Louisiana | Amite County, Mississippi | 0.96 | 82.3 (8%) |
| McDonald County, Missouri | Barry County, Missouri | 0.96 | 83.9 (8%) |
| Benton County, Missouri | Camden County, Missouri | 0.96 | 83.8 (8%) |
| Dunklin County, Missouri | Greene County, Arkansas | 0.97 | 98.4 (8%) |
| Bedford County, Tennessee | Moore County, Tennessee | 0.95 | 69.1 (8%) |
| Liberty County, Texas | Jefferson County, Texas | 0.96 | 82.3 (8%) |
| Boone County, West Virginia | Kanawha County, West Virginia | 0.98 | 109.6 (8%) |
| Walker County, Alabama | Jefferson County, Alabama | 0.97 | 103.5 (7.5%) |
| Bacon County, Georgia | Coffee County, Georgia | 0.98 | 90.4 (7.5%) |
| Murray County, Georgia | Gilmer County, Georgia | 0.97 | 93.1 (7.5%) |
| Accomack County, Virginia | Worcester County, Maryland | 0.97 | 78.1 (7.5%) |
| Dunklin County, Missouri | Craighead County, Arkansas | 0.96 | 99 (7.5%) |
| Wayne County, Missouri | Reynolds County, Missouri | 0.96 | 94.1 (7.5%) |
| Dewey County, South Dakota | Ziebach County, South Dakota | 0.96 | 79.1 (7.5%) |
| Refugio County, Texas | Bee County, Texas | 0.95 | 61.2 (7.5%) |
| Dallam County, Texas | Union County, New Mexico | 0.95 | 56.6 (7.5%) |
| San Patricio County, Texas | Live Oak County, Texas | 0.96 | 62.6 (7.5%) |
| Peach County, Georgia | Taylor County, Georgia | 0.96 | 82.7 (7%) |
| Scott County, Indiana | Jefferson County, Indiana | 0.97 | 112.5 (7%) |
| Bienville Parish, Louisiana | Claiborne Parish, Louisiana | 0.95 | 83.2 (7%) |
| Caldwell Parish, Louisiana | Richland Parish, Louisiana | 0.97 | 104.8 (7%) |
| Marlboro County, South Carolina | Darlington County, South Carolina | 0.97 | 113.7 (7%) |
| Baca County, Colorado | Las Animas County, Colorado | 0.94 | 48.8 (7%) |
| Okeechobee County, Florida | Glades County, Florida | 0.95 | 78.1 (6.5%) |
| Bleckley County, Georgia | Pulaski County, Georgia | 0.96 | 76.4 (6.5%) |
| Peach County, Georgia | Macon County, Georgia | 0.97 | 83.2 (6.5%) |
| Shoshone County, Idaho | Clearwater County, Idaho | 0.95 | 80.7 (6.5%) |
| Scott County, Indiana | Clark County, Indiana | 0.97 | 113.1 (6.5%) |
| Starke County, Indiana | LaPorte County, Indiana | 0.96 | 95.8 (6.5%) |
| Gallatin County, Illinois | Posey County, Indiana | 0.96 | 89.4 (6.5%) |
| Union County, Kentucky | Posey County, Indiana | 0.97 | 94.9 (6.5%) |
| Breathitt County, Kentucky | Magoffin County, Kentucky | 0.98 | 135 (6.5%) |
| Franklin County, Tennessee | Moore County, Tennessee | 0.94 | 63.3 (6.5%) |
| Scott County, Tennessee | Morgan County, Tennessee | 0.97 | 117.3 (6.5%) |
| Hopewell City, Virginia | Prince George County, Virginia | 0.98 | 94.7 (6.5%) |

| | | | |
|---|---|---|---|
| Webster County, West Virginia | Randolph County, West Virginia | 0.96 | 83 (6.5%) |
| Gordon County, Georgia | Gilmer County, Georgia | 0.95 | 81.9 (6%) |
| Macon County, Georgia | Dooly County, Georgia | 0.95 | 66.7 (6%) |
| Elko County, Nevada | Cassia County, Idaho | 0.93 | 56.2 (6%) |
| Pottawattamie County, Iowa | Shelby County, Iowa | 0.94 | 66.2 (6%) |
| Lee County, Kentucky | Jackson County, Kentucky | 0.97 | 129.7 (6%) |
| Grenada County, Mississippi | Carroll County, Mississippi | 0.95 | 92.5 (6%) |
| Polk County, Tennessee | Cherokee County, North Carolina | 0.96 | 101.9 (6%) |
| Sheridan County, Montana | Williams County, North Dakota | 0.94 | 62.4 (6%) |
| Polk County, Tennessee | Bradley County, Tennessee | 0.96 | 101.9 (6%) |
| Grainger County, Tennessee | Jefferson County, Tennessee | 0.95 | 93.2 (6%) |
| Anderson County, Texas | Freestone County, Texas | 0.96 | 99 (6%) |
| Pasco County, Florida | Sumter County, Florida | 0.94 | 68 (5.5%) |
| Breathitt County, Kentucky | Knott County, Kentucky | 0.97 | 136.4 (5.5%) |
| Powell County, Kentucky | Estill County, Kentucky | 0.97 | 142.5 (5.5%) |
| Pulaski County, Missouri | Camden County, Missouri | 0.94 | 82.5 (5.5%) |
| Union County, Illinois | Perry County, Missouri | 0.94 | 78.9 (5.5%) |
| Mohave County, Arizona | La Paz County, Arizona | 0.95 | 70.2 (5.5%) |
| San Bernardino County, California | La Paz County, Arizona | 0.92 | 41.4 (5.5%) |
| San Patricio County, Texas | Bee County, Texas | 0.94 | 64 (5.5%) |
| Wise County, Virginia | Norton City, Virginia | 0.94 | 96.8 (5.5%) |
| Lawrence County, Kentucky | Wayne County, West Virginia | 0.96 | 115.7 (5.5%) |
| Walker County, Alabama | Tuscaloosa County, Alabama | 0.95 | 106.3 (5%) |
| Scott County, Indiana | Washington County, Indiana | 0.97 | 114.9 (5%) |
| Comanche County, Kansas | Woods County, Oklahoma | 0.94 | 65.6 (5%) |
| Mississippi County, Missouri | Hickman County, Kentucky | 0.95 | 97.2 (5%) |
| Pike County, Kentucky | Dickenson County, Virginia | 0.95 | 112.7 (5%) |
| Lee County, Mississippi | Chickasaw County, Mississippi | 0.95 | 101.4 (5%) |
| Washington County, Mississippi | Issaquena County, Mississippi | 0.95 | 76.3 (5%) |
| Lake County, Tennessee | Dyer County, Tennessee | 0.96 | 112.6 (5%) |
| Trousdale County, Tennessee | Wilson County, Tennessee | 0.95 | 97.3 (5%) |
| Citrus County, Florida | Sumter County, Florida | 0.94 | 69 (4.5%) |
| Lincoln County, Georgia | McCormick County, South Carolina | 0.94 | 70.1 (4.5%) |
| Sumter County, Georgia | Dooly County, Georgia | 0.93 | 64.4 (4.5%) |
| Ben Hill County, Georgia | Wilcox County, Georgia | 0.94 | 90.1 (4.5%) |
| Saline County, Illinois | Pope County, Illinois | 0.94 | 91.6 (4.5%) |
| Scott County, Indiana | Jackson County, Indiana | 0.95 | 115.5 (4.5%) |
| Minnehaha County, South Dakota | Lyon County, Iowa | 0.92 | 56.3 (4.5%) |
| Estill County, Kentucky | Clark County, Kentucky | 0.95 | 114 (4.5%) |
| Breathitt County, Kentucky | Wolfe County, Kentucky | 0.96 | 137.9 (4.5%) |
| Perry County, Pennsylvania | Juniata County, Pennsylvania | 0.93 | 63.9 (4.5%) |

| | | | |
|---|---|---|---|
| Cheatham County, Tennessee | Williamson County, Tennessee | 0.94 | 91.8 (4.5%) |
| Walker County, Georgia | Hamilton County, Tennessee | 0.95 | 91 (4.5%) |
| Polk County, Texas | Angelina County, Texas | 0.95 | 86.8 (4.5%) |
| Gloucester County, Virginia | James City County, Virginia | 0.94 | 73.9 (4.5%) |
| Jackson County, Alabama | Franklin County, Tennessee | 0.94 | 90.5 (4%) |
| Levy County, Florida | Marion County, Florida | 0.94 | 89.5 (4%) |
| Putnam County, Florida | Flagler County, Florida | 0.93 | 87.1 (4%) |
| Jeff Davis County, Georgia | Coffee County, Georgia | 0.95 | 91.9 (4%) |
| Pottawatomie County, Kansas | Nemaha County, Kansas | 0.93 | 54.3 (4%) |
| Elk County, Kansas | Wilson County, Kansas | 0.94 | 74.9 (4%) |
| Leslie County, Kentucky | Clay County, Kentucky | 0.95 | 135.2 (4%) |
| Beauregard Parish, Louisiana | Allen Parish, Louisiana | 0.93 | 74 (4%) |
| Grenada County, Mississippi | Montgomery County, Mississippi | 0.94 | 94.4 (4%) |
| Pulaski County, Missouri | Texas County, Missouri | 0.94 | 83.8 (4%) |
| Bergen County, New Jersey | Bronx County, New York | 0.92 | 37.5 (4%) |
| Westchester County, New York | Bronx County, New York | 0.92 | 36.5 (4%) |
| Wibaux County, Montana | Golden Valley County, North Dakota | 0.92 | 50 (4%) |
| Mohave County, Arizona | Coconino County, Arizona | 0.93 | 71.3 (4%) |
| Humboldt County, Nevada | Malheur County, Oregon | 0.92 | 66.5 (4%) |
| Clay County, South Dakota | Cedar County, Nebraska | 0.93 | 50.1 (4%) |
| Lake County, Tennessee | Obion County, Tennessee | 0.95 | 113.8 (4%) |
| Phillips County, Arkansas | Desha County, Arkansas | 0.94 | 94.1 (4%) |
| Union County, Arkansas | Claiborne Parish, Louisiana | 0.94 | 85.2 (4%) |
| Rockingham County, Virginia | Pendleton County, West Virginia | 0.92 | 45.2 (4%) |
| Mingo County, West Virginia | Wayne County, West Virginia | 0.93 | 112 (4%) |
| Baca County, Colorado | Bent County, Colorado | 0.92 | 50.4 (4%) |
| Walker County, Alabama | Blount County, Alabama | 0.94 | 108 (3.5%) |
| Levy County, Florida | Citrus County, Florida | 0.93 | 89.9 (3.5%) |
| Decatur County, Georgia | Baker County, Georgia | 0.94 | 80 (3.5%) |
| Stephens County, Georgia | Oconee County, South Carolina | 0.94 | 85.9 (3.5%) |
| Franklin County, Kentucky | Shelby County, Kentucky | 0.93 | 89.7 (3.5%) |
| Harlan County, Kentucky | Lee County, Virginia | 0.95 | 129.3 (3.5%) |
| Marion County, Kentucky | Washington County, Kentucky | 0.94 | 92.3 (3.5%) |
| Yazoo County, Mississippi | Issaquena County, Mississippi | 0.93 | 71.3 (3.5%) |
| Yalobusha County, Mississippi | Tallahatchie County, Mississippi | 0.94 | 97.2 (3.5%) |
| Macon County, Tennessee | Sumner County, Tennessee | 0.94 | 96.5 (3.5%) |
| Monroe County, Arkansas | Saint Francis County, Arkansas | 0.94 | 95.8 (3.5%) |
| Adams County, Wisconsin | Wood County, Wisconsin | 0.93 | 77.2 (3.5%) |
| Okeechobee County, Florida | Highlands County, Florida | 0.93 | 81 (3%) |
| Wilcox County, Georgia | Dooly County, Georgia | 0.92 | 67.8 (3%) |
| Harlan County, Kentucky | Bell County, Kentucky | 0.94 | 129.9 (3%) |

| | | | |
|---|---|---|---|
| Daviess County, Kentucky | Warrick County, Indiana | 0.92 | 88 (3%) |
| Lawrence County, Kentucky | Elliott County, Kentucky | 0.93 | 118.8 (3%) |
| Whitley County, Kentucky | Laurel County, Kentucky | 0.93 | 120.2 (3%) |
| Perry County, Kentucky | Letcher County, Kentucky | 0.95 | 144.5 (3%) |
| LaSalle Parish, Louisiana | Rapides Parish, Louisiana | 0.92 | 81.9 (3%) |
| Iron County, Missouri | Dent County, Missouri | 0.92 | 97.2 (3%) |
| Iron County, Missouri | Reynolds County, Missouri | 0.93 | 97.2 (3%) |
| Jefferson County, Missouri | Monroe County, Illinois | 0.92 | 80.5 (3%) |
| Dewey County, South Dakota | Corson County, South Dakota | 0.93 | 83 (3%) |
| Marion County, Tennessee | Franklin County, Tennessee | 0.93 | 91.9 (3%) |
| Loving County, Texas | Reeves County, Texas | 0.93 | 76.6 (3%) |
| Polk County, Texas | Tyler County, Texas | 0.92 | 88.1 (3%) |
| Russell County, Alabama | Bullock County, Alabama | 0.92 | 86 (2.5%) |
| Murray County, Georgia | Bradley County, Tennessee | 0.93 | 98.1 (2.5%) |
| Scott County, Indiana | Jennings County, Indiana | 0.93 | 117.9 (2.5%) |
| Casey County, Kentucky | Adair County, Kentucky | 0.93 | 105.5 (2.5%) |
| Lawrence County, Kentucky | Carter County, Kentucky | 0.92 | 119.4 (2.5%) |
| Caldwell Parish, Louisiana | Catahoula Parish, Louisiana | 0.92 | 109.9 (2.5%) |
| Alcorn County, Mississippi | Tippah County, Mississippi | 0.92 | 83.9 (2.5%) |
| Cocke County, Tennessee | Haywood County, North Carolina | 0.92 | 96.5 (2.5%) |
| Marlboro County, South Carolina | Scotland County, North Carolina | 0.93 | 119.3 (2.5%) |
| Morgan County, Tennessee | Cumberland County, Tennessee | 0.93 | 88.8 (2.5%) |
| Monroe County, Arkansas | Lee County, Arkansas | 0.92 | 96.8 (2.5%) |
| Harrison County, West Virginia | Barbour County, West Virginia | 0.91 | 79.1 (2.5%) |
| Randolph County, West Virginia | Grant County, West Virginia | 0.91 | 58.1 (2.5%) |
| Hernando County, Florida | Sumter County, Florida | 0.92 | 72.4 (2%) |
| Polk County, Tennessee | Fannin County, Georgia | 0.92 | 106.2 (2%) |
| Monroe County, Iowa | Appanoose County, Iowa | 0.91 | 72.5 (2%) |
| East Carroll Parish, Louisiana | Issaquena County, Mississippi | 0.92 | 89.6 (2%) |
| Accomack County, Virginia | Somerset County, Maryland | 0.93 | 82.7 (2%) |
| Webster County, Mississippi | Oktibbeha County, Mississippi | 0.92 | 89.1 (2%) |
| Saint Francois County, Missouri | Perry County, Missouri | 0.92 | 93.7 (2%) |
| Fallon County, Montana | Golden Valley County, North Dakota | 0.91 | 51.8 (2%) |
| Graham County, Arizona | Apache County, Arizona | 0.91 | 42.8 (2%) |
| Dewey County, Oklahoma | Woodward County, Oklahoma | 0.92 | 72.8 (2%) |
| McDonald County, Missouri | Benton County, Arkansas | 0.92 | 89.4 (2%) |
| Lincoln County, West Virginia | Putnam County, West Virginia | 0.94 | 104.6 (2%) |
| Clinch County, Georgia | Echols County, Georgia | 0.91 | 99.9 (1.5%) |
| Peach County, Georgia | Houston County, Georgia | 0.91 | 87.6 (1.5%) |
| Clinch County, Georgia | Ware County, Georgia | 0.90 | 99.9 (1.5%) |
| Jay County, Indiana | Adams County, Indiana | 0.90 | 74 (1.5%) |

| | | | |
|---|---|---|---|
| Clay County, Kentucky | Laurel County, Kentucky | 0.93 | 121 (1.5%) |
| Hart County, Kentucky | Green County, Kentucky | 0.90 | 105 (1.5%) |
| Letcher County, Kentucky | Wise County, Virginia | 0.93 | 118.4 (1.5%) |
| Estill County, Kentucky | Madison County, Kentucky | 0.91 | 117.6 (1.5%) |
| Saint Bernard Parish, Louisiana | Plaquemines Parish, Louisiana | 0.90 | 93.2 (1.5%) |
| Jefferson County, Missouri | Sainte Genevieve County, Missouri | 0.90 | 81.7 (1.5%) |
| Nye County, Nevada | White Pine County, Nevada | 0.91 | 85.7 (1.5%) |
| Athens County, Ohio | Morgan County, Ohio | 0.90 | 73.3 (1.5%) |
| Bristol City, Virginia | Sullivan County, Tennessee | 0.91 | 96.6 (1.5%) |
| Coleman County, Texas | Concho County, Texas | 0.91 | 72.8 (1.5%) |
| Leon County, Texas | Madison County, Texas | 0.90 | 78.6 (1.5%) |
| Hopewell City, Virginia | Chesterfield County, Virginia | 0.92 | 99.8 (1.5%) |
| Lee County, Virginia | Scott County, Virginia | 0.90 | 98.2 (1.5%) |
| Martinsville City, Virginia | Henry County, Virginia | 0.91 | 78.9 (1.5%) |
| Yuba County, California | Sierra County, California | 0.91 | 71.4 (1.5%) |
| Pike County, Alabama | Bullock County, Alabama | 0.90 | 74.2 (1%) |
| Bryan County, Georgia | Bulloch County, Georgia | 0.90 | 83.7 (1%) |
| Bryan County, Georgia | Liberty County, Georgia | 0.91 | 83.7 (1%) |
| Baker County, Florida | Charlton County, Georgia | 0.91 | 84.5 (1%) |
| Toombs County, Georgia | Emanuel County, Georgia | 0.92 | 90.3 (1%) |
| Alexander County, Illinois | Cape Girardeau County, Missouri | 0.90 | 97 (1%) |
| Daviess County, Kentucky | Spencer County, Indiana | 0.90 | 89.8 (1%) |
| Hardin County, Illinois | Livingston County, Kentucky | 0.92 | 104.9 (1%) |
| Wolfe County, Kentucky | Menifee County, Kentucky | 0.91 | 117 (1%) |
| Webster County, Mississippi | Clay County, Mississippi | 0.92 | 90 (1%) |
| Scott County, Missouri | Cape Girardeau County, Missouri | 0.90 | 90.2 (1%) |
| Fallon County, Montana | Slope County, North Dakota | 0.91 | 52.4 (1%) |
| Marlboro County, South Carolina | Dillon County, South Carolina | 0.92 | 121.1 (1%) |
| Carroll County, Tennessee | Madison County, Tennessee | 0.91 | 92.5 (1%) |
| Rhea County, Tennessee | Bledsoe County, Tennessee | 0.91 | 89.1 (1%) |
| Warren County, Tennessee | Van Buren County, Tennessee | 0.90 | 83.5 (1%) |
| Wheeler County, Texas | Roger Mills County, Oklahoma | 0.91 | 66.4 (1%) |
| Grand County, Utah | Emery County, Utah | 0.90 | 53.5 (1%) |
| Montrose County, Colorado | San Juan County, Utah | 0.90 | 45.9 (1%) |
| Cleveland County, Arkansas | Lincoln County, Arkansas | 0.90 | 74.7 (1%) |
| Franklin County, Alabama | Marion County, Alabama | 0.90 | 99.5 (0.5%) |
| Monroe County, Florida | Miami-Dade County, Florida | 0.91 | 56.8 (0.5%) |
| Pike County, Georgia | Upson County, Georgia | 0.90 | 84.5 (0.5%) |
| Bacon County, Georgia | Ware County, Georgia | 0.90 | 97.3 (0.5%) |
| Schuyler County, Illinois | Brown County, Illinois | 0.91 | 66.7 (0.5%) |
| Orange County, Indiana | Dubois County, Indiana | 0.90 | 82.5 (0.5%) |

| | | | |
|---|---|---|---|
| White County, Illinois | Gibson County, Indiana | 0.90 | 80.4 (0.5%) |
| Geary County, Kansas | Clay County, Kansas | 0.90 | 78.3 (0.5%) |
| Butler County, Kansas | Marion County, Kansas | 0.90 | 67.8 (0.5%) |
| Lee County, Kentucky | Estill County, Kentucky | 0.90 | 137.3 (0.5%) |
| McCreary County, Kentucky | Campbell County, Tennessee | 0.90 | 142.8 (0.5%) |
| Floyd County, Kentucky | Pike County, Kentucky | 0.90 | 132.2 (0.5%) |
| Mercer County, Kentucky | Washington County, Kentucky | 0.91 | 93.6 (0.5%) |
| Grant Parish, Louisiana | Rapides Parish, Louisiana | 0.91 | 87.4 (0.5%) |
| Saint Bernard Parish, Louisiana | Orleans Parish, Louisiana | 0.90 | 94.2 (0.5%) |
| Daviess County, Missouri | DeKalb County, Missouri | 0.90 | 71.5 (0.5%) |
| San Juan County, New Mexico | McKinley County, New Mexico | 0.90 | 38.6 (0.5%) |
| Sierra County, New Mexico | Catron County, New Mexico | 0.90 | 57.9 (0.5%) |
| Towner County, North Dakota | Benson County, North Dakota | 0.91 | 48.1 (0.5%) |
| Brown County, South Dakota | McPherson County, South Dakota | 0.90 | 52.8 (0.5%) |
| Fall River County, South Dakota | Sioux County, Nebraska | 0.90 | 66.8 (0.5%) |
| McNairy County, Tennessee | Chester County, Tennessee | 0.91 | 95.3 (0.5%) |
| Putnam County, Tennessee | Cumberland County, Tennessee | 0.90 | 80 (0.5%) |
| Liberty County, Texas | Harris County, Texas | 0.91 | 89 (0.5%) |
| Shackelford County, Texas | Jones County, Texas | 0.92 | 69.1 (0.5%) |
| Trinity County, Texas | Angelina County, Texas | 0.90 | 86.8 (0.5%) |
| Warren County, Virginia | Page County, Virginia | 0.90 | 75.5 (0.5%) |
| Lincoln County, West Virginia | Cabell County, West Virginia | 0.91 | 106.2 (0.5%) |
| Taylor County, West Virginia | Preston County, West Virginia | 0.90 | 71.7 (0.5%) |

Table S1. Reported spatial disparities and Residual Disparity Elimination Targets (RDETs) between neighboring counties from boundary analysis of 2014 US county-level tracheal, bronchus, and lung cancer mortality rates. The RDET represents the minimum reduction in mortality rate at which the higher residual mortality county would not be classified as a disparity with the neighboring county. Boundary analysis was conducted using a Bayesian spatial regression model and posterior samples were obtained using a trained neural posterior estimator.

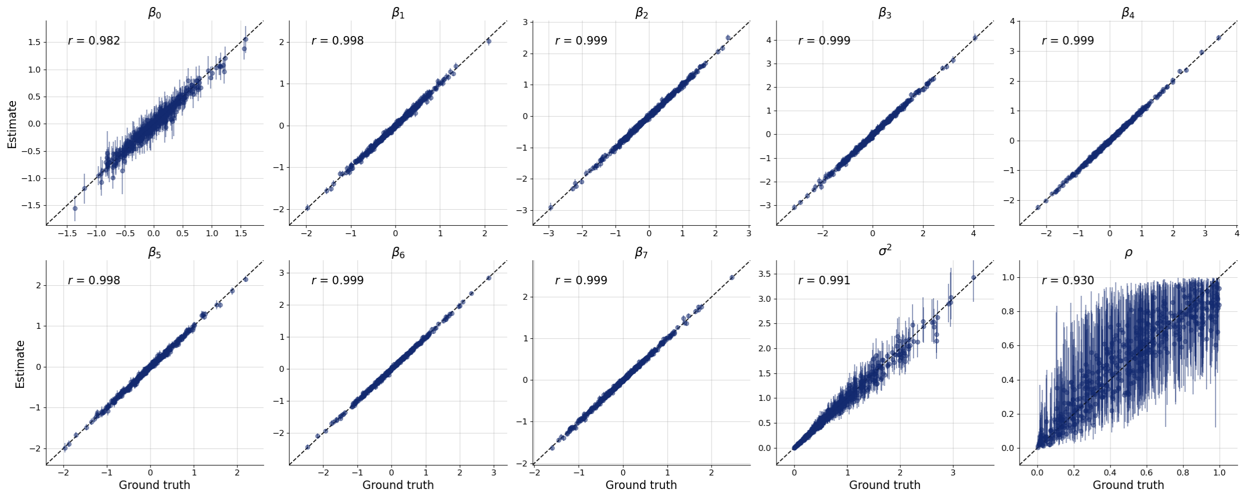


FIGURE S3. Recovery plot of credible intervals for regression coefficient and variance parameters generated by neural posterior estimation network trained without an auxiliary network versus true values generated from the prior across 500 simulated datasets.

S. T. Radev, U. K. Mertens, A. Voss, L. Ardizzone, and U. Köthe. BayesFlow: Learning Complex Stochastic Models With Invertible Neural Networks. *IEEE Transactions on Neural Networks and Learning Systems*, 33(4):1452–1466, Apr. 2022. ISSN 2162-2388. doi: 10.1109/TNNLS.2020.3042395. URL `https://ieeexplore.ieee.org/document/9298920`.

S. Talts, M. Betancourt, D. Simpson, A. Vehtari, and A. Gelman. Validating Bayesian Inference Algorithms with Simulation-Based Calibration, 2018. URL `https://arxiv.org/abs/1804.06788`. Version Number: 2.

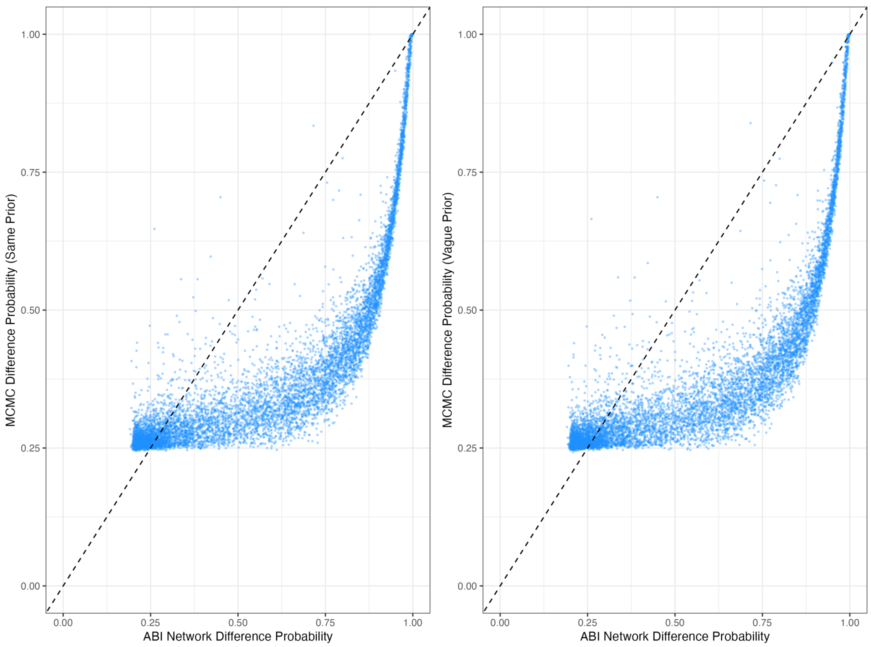


FIGURE S4. Neighboring county-pairs' residual difference probabilities obtained using Markov Chain Monte Carlo (MCMC) with an equivalent prior (left) and non-informative prior (right) versus difference probabilities obtained using a neural posterior estimation network trained without an auxiliary network. The 45-degree reference line (dotted) denotes perfect equality.
